# The influence of biological, epidemiological, and treatment factors on the establishment and spread of drug-resistant *Plasmodium falciparum*

**DOI:** 10.1101/2022.02.05.22270500

**Authors:** Thiery Masserey, Tamsin Lee, Monica Golumbeanu, Andrew J Shattock, Sherrie L Kelly, Ian M Hastings, Melissa A Penny

**Author notes:** **Corresponding author:** Name: Melissa Penny, Institution: Swiss Tropical and Public Health Institute, Basel, Switzerland. University of Basel, Basel, Switzerland.

## Abstract

The effectiveness of artemisinin-based combination therapies (ACTs) to treat *Plasmodium falciparum* malaria is threatened by resistance. The complex interplay between sources of selective pressure – treatment properties, biological factors, transmission intensity, and access to treatment – obscures understanding how, when, and why resistance establishes and spreads across different locations. We developed a disease modelling approach with emulator-based global sensitivity analysis to systematically quantify which of these factors drive establishment and spread of drug resistance. Drug resistance was more likely to evolve in low transmission settings due to the lower levels of (i) immunity and (ii) within-host competition between genotypes. Spread of parasites resistant to artemisinin partner drugs depended on the period of low drug concentration (known as the selection window). Spread of partial artemisinin resistance was slowed with prolonged parasite exposure to artemisinin derivatives and accelerated when the parasite was also resistant to the partner drug. Thus, to slow the spread of partial artemisinin resistance, molecular surveillance should be supported to detect resistance to partner drugs and to change ACTs accordingly. Furthermore, implementing more sustainable artemisinin-based therapies will require extending parasite exposure to artemisinin derivatives, and mitigating the selection windows of partner drugs, which could be achieved by including an additional long-acting drug.

**Impact Statement:** Detailed models of malaria and treatment dynamics were combined with emulator-based global sensitivity analysis to elucidate how the interplay of drug properties, infection biology, and epidemiological dynamics drives evolution of resistance to artemisinin-based combination therapies. The results identify mitigation strategies.

## Introduction

Malaria remains a global health priority [1]. The World Health Organization (WHO) recommends several artemisinin-based combination therapies (ACTs) to treat uncomplicated *Plasmodium falciparum* malaria [2]. ACTs combine a short-acting artemisinin derivative to rapidly reduce parasitaemia during the first three days of treatment and a long-acting partner drug to eliminate remaining parasites [2]. These drug combinations are intended to delay the evolution of drug resistance, which has frequently occurred under monotherapy treatment [3–6]. However, parasites partially resistant to artemisinin have emerged in the Greater Mekong Subregion (GMS) and, more recently, in Rwanda, Uganda, Guyana, and Papua New Guinea despite the use of ACTs [2, 7–11]. Partial artemisinin resistance leads to slower parasite clearance following treatment with ACTs, but not necessarily to treatment failure [2]. However, high rates of treatment failure have been observed in the GMS due to parasites being less sensitive to artemisinin derivatives and their partner drugs [2]. To prevent the evolution of drug-resistant parasites and to preserve the efficacy of ACTs or triple combination therapies (TACT, including a second long-acting drug) now being tested [12], it is essential to understand which factors drive this process.

The evolution of drug resistance follows a three-step process of mutation, establishment, and spread. First, mutations conferring drug resistance emerge in the population at a rate that depends on multiple factors, such as organism mutation and migration rates [13, 14]. Second, establishment is a highly stochastic step as the parasite with the drug-resistant mutation needs to infect other hosts [13–16]. The resistant strain establishes in the population once its frequency is high enough to minimise its risk of stochastic extinction [13–16]. Several forces influence the establishment of mutations. In settings with higher heterogeneity of parasite reproductive success, establishment of mutations is less likely because the effects of stochasticity are more substantial [13, 15–17]. This heterogeneity depends on the level of transmission and health system strength [13, 15–18]. In addition, the more selection favours the resistant strain, the more likely it is to establish [13, 15–17]. The strength of selection depends on many factors, such as the parasite and human biology, the transmission setting, drug properties, and health system strength [5, 19–24]. Third, resistance spreads through a region after a resistant mutation has become established. The mutation spreads at a rate that depends on the strength of selection [13, 16].

It is not fully understood how factors intrinsic to the transmission setting, health system, human and parasite biology, and drug properties interact to influence the establishment and spread of drug-resistant parasites. Mathematical models of infectious disease have not previously been used to systematically assess the joint influence of multiple factors on the establishment and spread of drug resistance, e.g. [23, 25–37]. Simple models, based on the Ross and MacDonald model [38, 39], have considered specific components of the epidemiology of resistance and, therefore, are not sophisticated enough to answer questions on how factors have jointly impacted establishment and spread of drug resistance [26, 30–33, 36, 37]. Most models have investigated specific transmission scenarios and questions, such as how within-host competition between parasites influences development of drug resistance [25, 28, 35], and did not systematically assess the impact of assumptions used on their results. Consequently, previous studies have not systematically compared the influence of multiple drivers, nor assessed how their influence varies under different transmission settings or health system strengths.

In addition, most models have made simplifications concerning drug action and consequences of partial resistance. They have not explicitly modelled the pharmacokinetics and pharmacodynamics of the drugs and have assumed that resistant parasites are fully resistant to the drugs. Parasites partially resistant to artemisinin exhibit an extended ring-stage during which they are not sensitive to artemisinin, however, parasites remain sensitive to artemisinin during other stages [40–44]. In addition, parasites resistant to partner drugs have an increased minimum inhibitory concentration (MIC), meaning that they are not sensitive to low drug concentrations but remain susceptible to high concentrations of partner drugs [45–47]. Consequently, many models have ignored the residual effect of drugs on resistant parasites and have not investigated the influence of the degree of resistance and drug proprieties on the establishment and spread of drug resistance. Models that have explicitly considered drug action have focused on specific questions such as how half-life impacts the spread of resistance or how resistance to the partner drug influences evolution of artemisinin resistance [48, 49]. However, they did not investigate how the impact of drug proprieties and the degree of resistance interact with other biological, transmission, and health system factors.

In this study, we developed a disease model with an emulator-based approach to quantify the influence of factors intrinsic to the biology of the parasite and human, the transmission setting, the health system strength, and the drug properties on the establishment and spread of drug-resistant parasites. Our approach is based on a detailed individual-based malaria model, OpenMalaria (https://github.com/SwissTPH/openmalaria/wiki), that includes a mechanistic within-host model (based on [50]). We first adapted our model, OpenMalaria, to explicitly include mechanistic drug action models at the individual level (as a one, two, or three-compartment pharmacokinetic model with a pharmacodynamics component of parasite killing [51–54]) and to track multiple parasite genotypes to which we could assign fitness costs and drug susceptibility (i.e. pharmacodynamic) properties. We then built an emulator-based workflow to quantify, through a series of global sensitivity analyses, the influence of multiple factors on the establishment and spread of parasites having different degrees of resistance to artemisinin derivatives and/or their partner drugs when used in monotherapy and combination (as ACTs). Emulators are predictive models that can approximate the relationship between input and output parameters of complex models and can run much faster than complex models to perform global sensitivity analyses more efficiently [55]. OpenMalaria is a mechanistic model, so the observed dynamics at the population level (for example, the spread of resistant genotypes) emerges from the relationship between the different model components and their input parameters. These dynamics can only be understood and tested through extensive analyses as undertaken here. Identifying which factors (e.g. drug properties and/or setting characteristics) favour the evolution of resistance, enables us to identify drug properties or strategies to slow or mitigate resistance and guides the development and implementation of more sustainable therapies.

## Results

### Development of drug resistance

We investigated the establishment and spread of drug-resistant genotypes by varying the degrees of resistance for three different treatment profiles. The first treatment profile considered was a monotherapy using a short-acting drug referred to in this study as drug A. Drug A has a short half-life and a high killing efficacy, simulating artemisinin derivatives (Figure 1A and 1B). Patients received a daily dose of drug A for six days (see Methods). To mimic the mechanism of resistance to artemisinin derivatives, we assumed that genotypes resistant to drug A had lower maximum killing rates (Emax) than sensitive ones (Figure 1B) (see Methods). We defined the degree of resistance to drug A as the relative decrease of the Emax of the resistant genotype compared with the sensitive one. The second treatment profile was also a monotherapy but with a long-acting drug referred to in this study as drug B. Drug B has a longer half-life and a lower Emax than drug A, typical of partner drugs used for ACTs (such as mefloquine, piperaquine, and lumefantrine) (Figure 1A and 1B). Patients received a daily dose of drug B for three days (see Methods). We assumed that genotypes resistant to drug B had higher half-maximal effective concentrations (EC50) than sensitive ones (Figure 1B) (see Methods). We defined the degree of resistance to drug B as the relative increase of the EC50 of the resistant genotype compared with the sensitive genotype. The last treatment profile was a daily dose of a combination of drugs A and B for three days, simulating ACTs. In this last case, only the resistant genotype had some degree of resistance to drug A, but both the sensitive and resistant genotypes could have the same degree of resistance to drug B.

**Figure 1.**
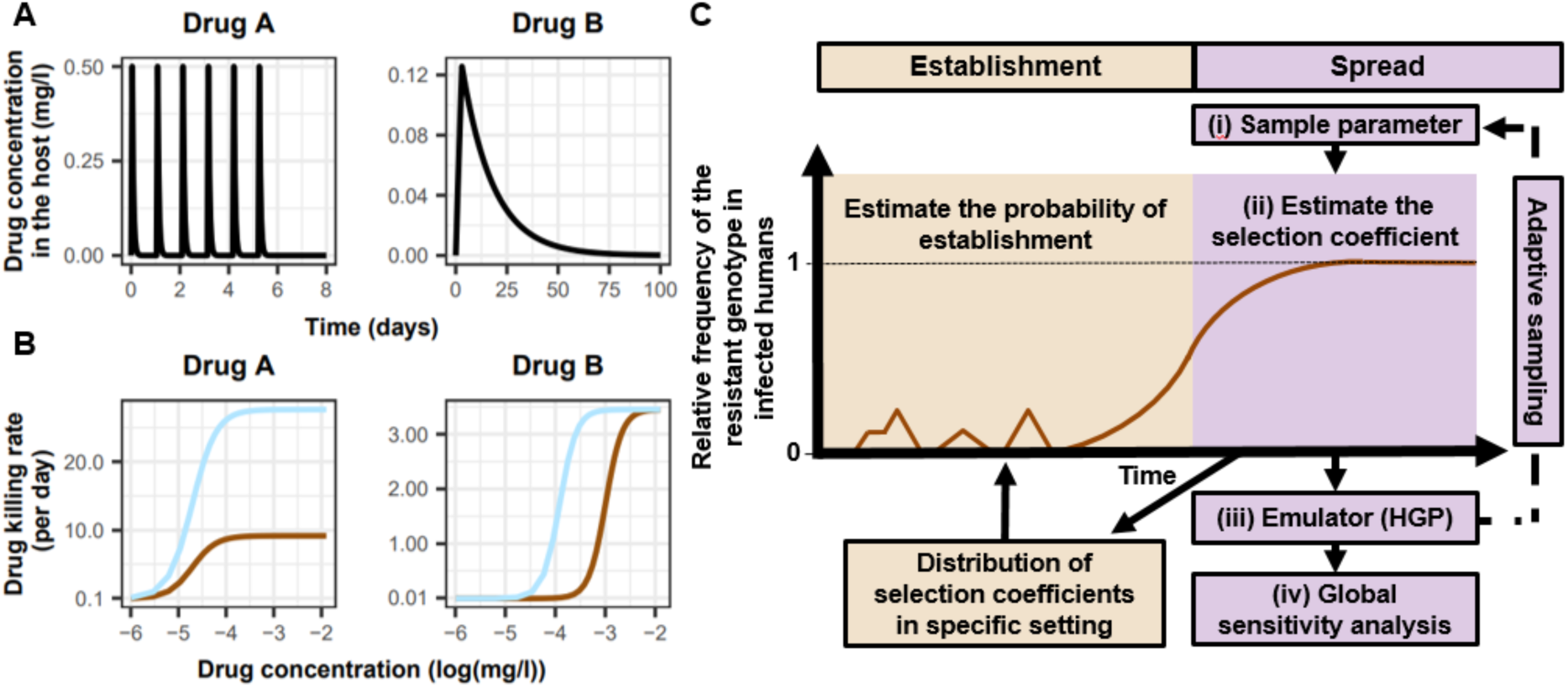
Overview of treatment profiles and the workflow. (**A**) Curves represent examples of the modelled within-host concentration (mg/l) of drugs A (short-acting like artemisinin derivatives) and B (long-acting like partner drugs of artemisinin) used in monotherapy. Patients received a daily dose of drug A for six days (see Methods). Patients received a daily dose of drug B for three days (see Methods). Drugs A and B used in combination (like ATCs) had the same respective profile as in monotherapy, but patients received a daily dosage of each drug over three days, as recommended by WHO for ACTs [56]. (**B**) Curves illustrate examples of the modelled relationship between the concentration (log(mg/l)) and the killing effect (per day) of drugs A and B on the resistant (brown) and sensitive genotypes (blue). For the use of drugs in combination, the resistant genotype was resistant to drug A, and both sensitive and resistant genotypes could have some degree of resistance to drug B. (**C**) The orange area highlights steps that evaluate how the probability of establishment of mutations with a specific selection coefficient varies under different settings. The purple area highlights the steps for assessing the influence of factors on the rate of spread (selection coefficient) of a resistant genotype through global sensitivity analysis. The brown curve represents an example of the relative frequency of the resistant genotype in infected humans. HGP: Heteroskedastic Gaussian Process.

Our analysis had two steps. First, we quantified the impact of factors listed in Table 1 on the spread of drug-resistant parasites through global sensitivity analyses using an emulator trained on our model simulations (Figure 1C, purple area, see Methods). For each simulation, we tracked a drug-sensitive genotype and a drug-resistant genotype, and we estimated the rate of spread using the selection coefficient, which measures the rate at which the logit of the resistant genotype frequency increases each parasite generation (see Methods, note that a selection coefficient below zero implies that resistance does not spread in the population) [16]. Then, we assessed the probability of establishment for a sub-set of resistant genotypes with known and positive selection coefficients to observe the relationship between selection coefficient and the probability of establishment (Figure 1C, orange area, see Methods). We could then extrapolate the probability of establishing any mutations with a known selection coefficient, which made the process more efficient since estimating the probability of establishment requires running many more stochastic realisations than estimating the selection coefficient due to the stochasticity of this step.

**Table 1.** Potential drivers of the spread of drug resistance. List of factors and their parameter ranges investigated in the global sensitivity analyses of the spread of parasites resistant to each treatment profile. The parameter ranges were defined based on the literature as described in the Methods. Parameter ranges of drug A captured the parameter values of typical artemisinin derivatives (see Methods). The parameter ranges of drug B captured the parameter values of partner drugs of artemisinin derivatives such as mefloquine, piperaquine, and lumefantrine (see Methods). Note that the ratio Cmax/EC50 is not a direct input of the model, but we varied this ratio by varying the EC50 of the sensitive genotype and the drug dosage (which impacted the maximum drug concentration (Cmax)) (see Methods). A Latin Hypercube Sampling (LHS) algorithm was used to sample from the ranges [57].

| Component | Determinant | Definition | Parameter range |  |
| --- | --- | --- | --- | --- |
|  |  |  | Drug A | Drug B |
| Drug properties | Half-life | Time for the drug concentration to fall by 50% (days) | (0.035, 0.175) | (6, 22) |
|  | Emax | Maximum killing rate the drug can achieve (per day) | (27.5, 31.0) | (3.45, 5.00) |
|  | Cmax/EC50 | The ratio between the maximum drug concentration (Cmax) and the half-maximal effective concentrations (EC50) of the sensitive genotype. This calculated ratio captures the drug killing effect by capturing how high is the Cmax compared to the EC50. | (55.0–312.0) | (5.1–21.7) |
| Parasite biology | Degree of resistance | For drug A: relative decrease of the Emax of the resistant genotype compared with the sensitive one<br>For drug B: relative increase of the EC50 of the resistant genotype compared with the sensitive one (see Methods) | (1, 50) | (1, 20) |
|  | Fitness cost | Relative reduction of the resistant genotype multiplication rate within the human host compared to the sensitive one | (1.0, 1.1) |  |
| Transmission level | Entomological inoculation rate | Mean number of infective mosquito bites received by an individual during a year (inoculations per person per year) | (5, 500) |  |
| Health system | Level of access to treatment | The probability of symptomatic cases to receive treatment within two weeks from the onset of symptom onset (%) | (10, 80) |  |
|  | Diagnostic detection limit | Parasite density for which the probability of having a positive diagnostic test is 50% (parasites/μl) | (2, 50) |  |

### Key drivers of the spread of drug-resistant parasites

Under monotherapy, access to treatment and degree of resistance of a monotherapy were the main drivers of the spread of resistance (Figure 2A). For drugs A and B used as monotherapy, the selection coefficient increased with increasing access to treatment (the probability of symptomatic cases to receive treatment within two weeks from the onset of symptoms) (Figure S1). In addition, higher degrees of resistance of the resistant genotype to drug A (relative decrease in the resistant genotype Emax compared with the sensitive one) and B (relative increase in the resistant genotype EC50 compared with the sensitive one) promoted the spread of parasites resistant to drugs A and B, respectively (Figure S1).

**Figure 2.**
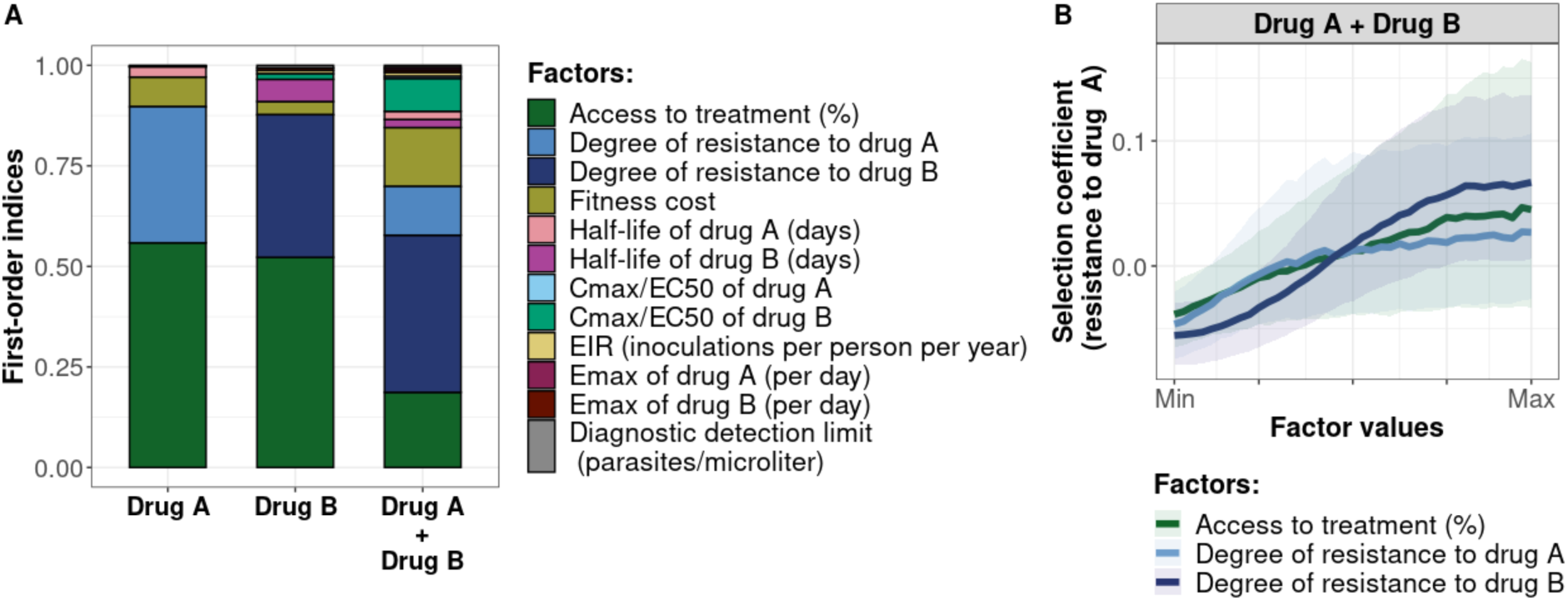
Influence of drug properties, fitness costs, resistance levels, transmission levels, and health system factors on predicted selection coefficients for three treatment profiles. (**A**) The first-order indices from our variance decomposition analysis indicate the level of importance of drug properties, fitness costs, resistance levels, transmission levels, access to treatment, and diagnostic limits in determining the spread of drug resistance. Indices are shown for each treatment profile in a non-seasonal setting with a population fully adherent to treatment. Selection coefficients are considered for drug A and drug B when each drug is used as monotherapy and for drug A when both drugs are used in combination. Definitions and ranges of parameters investigated are listed in Table 1. (**B**) Influence of factors on the selection coefficient of genotypes resistant to drug A in a population that used drugs A and B in combination. Curves and shaded areas represent the median and interquartile range of selection coefficients estimated during the global sensitivity analyses over the following parameter ranges: access to treatment (10−80%); the degree of resistance of the resistant genotype to drug A (1−50-fold reduction in Emax); and the degree of resistance of both sensitive and resistant genotypes to drug B (1−20-fold increase in EC50). A selection coefficient below zero implies that resistance does not spread in the population but is being lost due to its fitness costs. The transmission setting was non-seasonal and, all treated individuals were fully adherent to treatment.

When drugs A and B were used in combination, we assumed the resistant genotype had some degree of resistance to drug A, but both the sensitive and resistant genotypes could have some degree of resistance to drug B. In this case, the most important driver of spread was the degree of resistance of both genotypes to drug B (Figure 2A). The median selection coefficient was below zero when both genotypes were susceptible to drug B (the minimum degree of resistance to drug B) (Figure 2B), indicating that using an efficient partner drug can limit the spread of artemisinin resistance. The spread of parasites resistant to drug A was accelerated when parasites were also resistant to the partner drug, highlighting that resistance to the partner drug can facilitate the spread of artemisinin resistance. We further illustrated with concrete examples (Supplementary file 1: section 1.2) how the spread of partial resistance to drug A accelerates with higher degrees of resistance to drug B. These results further confirmed that resistance to partner drugs facilitates the spread of resistance to artemisinin, highlighting the importance of combining artemisinin derivatives with an efficient partner drug.

### Variation in the influence of factors across settings and degrees of resistance

We compared the effects of drug properties and fitness cost on the selection coefficients for a fixed set of degrees of resistance, transmission levels, seasonality patterns, treatment levels, and levels of adherence to treatment (percentage of treatment doses adhered by patients) (see the legend of Figure 3 for the values of each fixed factor). Across settings with a low access to treatment, we found that fitness cost had the largest influence on the selection coefficient (Figure S2-5). The fitness cost of a resistant genotype was defined as the relative decrease in the resistant genotype multiplication rate within an untreated human host compared with the sensitive genotype. Consequently, high fitness costs prevented the spread of resistance (Figure S2). At a high level of access to treatment, drug properties played a critical role in the spread of drug resistance, and their influence varied for each treatment profile as described below (Figure 3, Figure S3-5).

**Figure 3.**
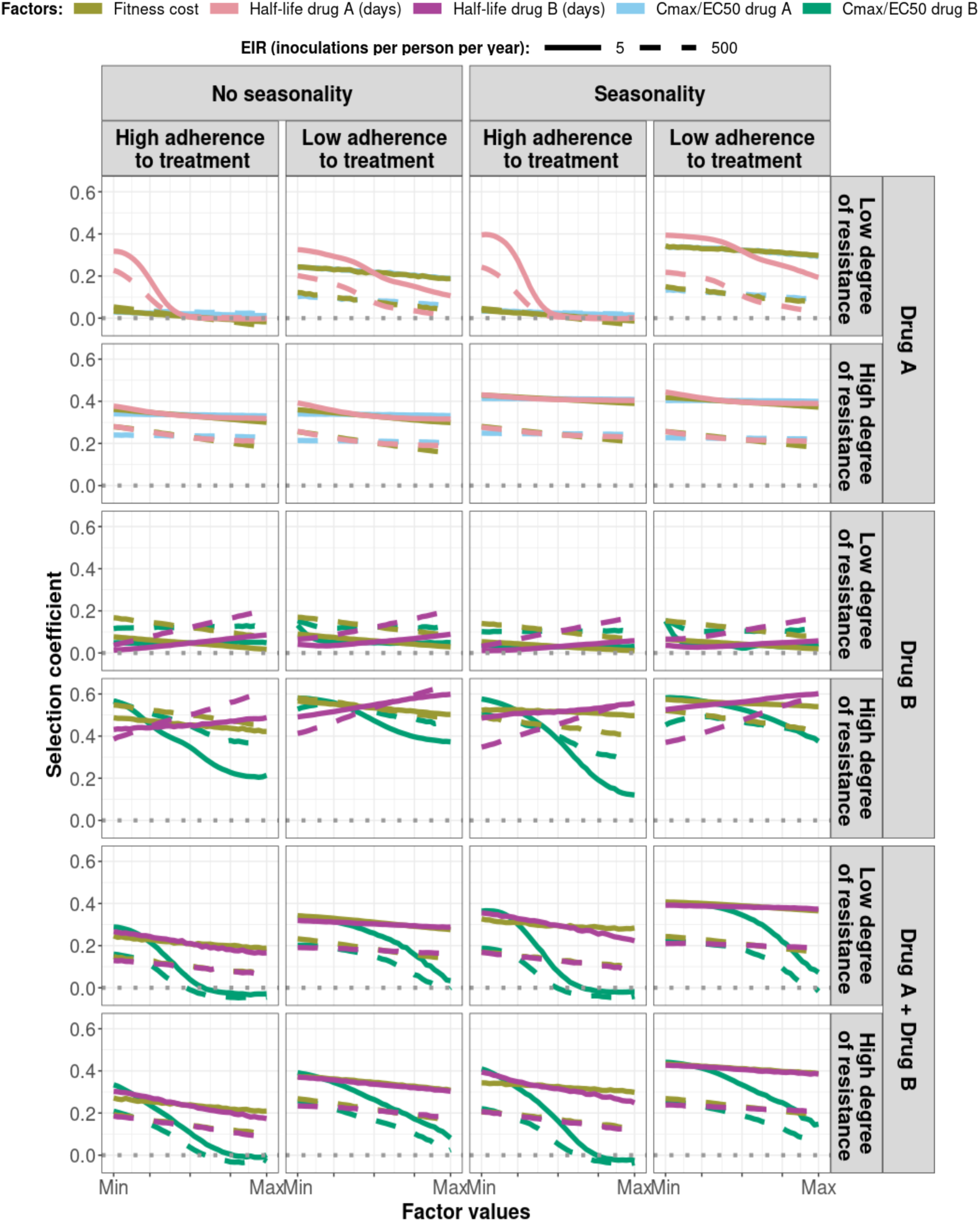
Magnitude and direction of effect of drug properties and fitness cost on predicted selection coefficients for low and high levels of transmission, degrees of drug resistance, treatment adherence, in seasonal or perennial settings with monotherapy or combination treatment. The curves represent median selection coefficients over the parameter ranges estimated in each setting that had high access to treatment (80%) and an entomological inoculation rate (EIR) of 5 (solid curves) or 500 (dashed curves) inoculations per person per year. Settings were varied in their seasonality pattern of transmission and level of adherence to treatment (67% (low) or 100% (high) of treatment doses adhered to by the population). For each treatment profile, results are shown for parasites with two different degrees of resistance; degree of resistance of 7 (low) and 18 (high) to drug A (Emax shift), 2.5 (low) and 10 (high) to drug B (EC50 shift), for the combination of drugs A and B, 7 (low) and 18 (high) to drug A and 10 to drug B. Parameter ranges are as follows: fitness cost (1.0−1.1, light green curves); drug A half-life (0.035−0.175 days, pink curves); drug B half-life (6−22 days, purple curves); Cmax/EC50 ratio for drug A (55.0−312.0, blue curves); Cmax/EC50 ratio for drug B (dark green curves) at a high level of adherence to treatment (5.4−21.7) and at a low level of adherence (4.0−16.2).

For drug A used as monotherapy, the half-life had the biggest influence on the rate of spread (Figure 3, Figure S3). A long half-life reduced the spread of resistant parasites by extending the period during which the drug killed partially resistant parasites (Figure 3). Furthermore, the spread of the resistant genotype was faster in populations with low adherence to treatment (Figure 3–Figure S6) because with fewer treatment doses, the parasite was exposed to the drug for a shorter time, leading to higher parasite survival. Overall, these results highlight that the time during which the parasite is exposed to artemisinin is a critical driver of the spread of partial artemisinin resistance.

For parasites resistant to drug B when used as monotherapy, the drug half-life also had an important influence on the selection coefficient (Figure 3, Figure S4). However, long half-lives were associated with large selection coefficients (Figure 3). Drugs with a long half-life have an extended period of low drug concentration in treated patients during which only resistant parasites can infect the host. This period of low drug concentration is called the selection window [49, 58]. These results confirm that the selection window plays a crucial role in the spread of resistance to long-acting drugs.

In addition, for parasites resistant to drug B as monotherapy, the drug killing rate captured by the ratio Cmax/EC50 had an important influence on the rate of spread in settings with low level of treatment adherence or a high degree of resistance (Figure 3, Figure S4). When we modelled a low level of treatment adherence or a high level of resistance, this ratio was reduced due to lower Cmax or higher EC50, respectively. Lower Cmax/EC50 ratios cause lower drug killing rates, and when this ratio was too low the spread of drug resistance was favoured. These results highlight the importance of treatment adherence to assure that the drug concentration is high enough to eliminate partially resistant genotypes and limit their spread.

When the genotype was resistant to drug A in a population that used drugs A and B in combination, factors related to drug B had the most influence on the selection coefficient (Figure 3, Figure S5). When the ratio Cmax/EC50 or the half-life of drug B was large, the killing effect of drug B on parasites resistant to drug A was higher, reducing their spread (Figure 3). In addition, the rate of spread rose when the level of adherence to treatment was low (Figure S6). These results highlight that the spread of partial resistance to artemisinin strongly depends on the capacity of the partner drug to kill them.

The influence of the transmission intensity (represented by EIR) and its seasonality on the selection coefficient varied by treatment profiles. When the parasite was resistant to drug A when used as monotherapy or in combination with drug B, selection coefficients were higher in settings with lower EIR (Figure S6). Two factors account for this trend. First, the selection of parasites resistant to drug A depends on the proportion of infections that are treated and can thus select for resistance. This proportion is higher at lower EIR due to the lower level of immunity (Figure S7) which makes infections more likely to be symptomatic and hence receive treatment. Second, there is a high multiplicity of infection in high transmission settings. The multiplicity of infection enhanced within-host competition between genotypes, which inhibit the multiplication of resistant parasites within hosts due to their fitness cost, and thus limit their spread. Similarly, the spread of resistant parasites was higher in the seasonal settings than in non-seasonal settings (Figure S6) due to the reduction of immunity levels and a decline in within-host competition between genotypes during the low transmission season of the seasonal settings. Overall, these results indicate that the spread of partial artemisinin resistance is faster in seasonal settings with low transmission levels.

However, for parasites resistant to drug B used in monotherapy, selection coefficients were higher in settings with a large EIR (Figure S6). This relationship arises because resistant parasites are more likely to emerge from the liver in high transmission settings during selection window (which select for resistant parasites). This is because the proportion of people treated (and thus with residual drug concentrations) is lower at lower EIR due to lower infection rates (Figure S7). Note that this trend was only valid for settings with high access to treatment. In settings with low access to treatment, we observe similar trends than for parasites resistant to drug A (Figure S8) since here, the impact of the selection window was more negligible. These results highlight that the selection window of the long-acting drug can change the interplay between the transmission setting and the spread of drug resistance.

### Probability of establishment of drug resistance and its key drivers

In each non-seasonal setting, we selected 10 different resistant genotypes having a known selection coefficient and quantified their probability of establishment (see Methods). By doing so, we evaluated the relationship between the selection coefficient and probability of establishment and assessed how this relationship varies across settings due to variation in the heterogeneity of parasite reproductive success.

As expected, the establishment of a mutation was more probable when its selection coefficient was high (Figure 4). For each treatment profile, the probability of establishment of mutations with similar selection coefficients was higher at low EIR than at high EIR (Figure 4), especially for mutations with a high selection coefficient. These results highlight that the heterogeneity in parasite reproductive success increases with the transmission level, causing more uncertainty in the establishment of mutations. Two factors increase the heterogeneity of parasite reproductive success in settings with a high EIR. First, in these settings, there is considerable variation in the number of independent infections carried by hosts, which are competing for reproductive success. This variation leads to more heterogeneity of parasite reproductive success and thus to less certainty that a parasite with an emerging mutation will replicate. Second, settings with a high EIR have a large variation in the level of individual immunity. Host immunity influences parasite reproductive success by reducing parasite growth within the human host. Therefore, in high transmission settings, the greater variation of immunity leads to higher heterogeneity of parasite reproductive success and a lower probability that an emerging mutation successfully replicate.

**Figure 4.**
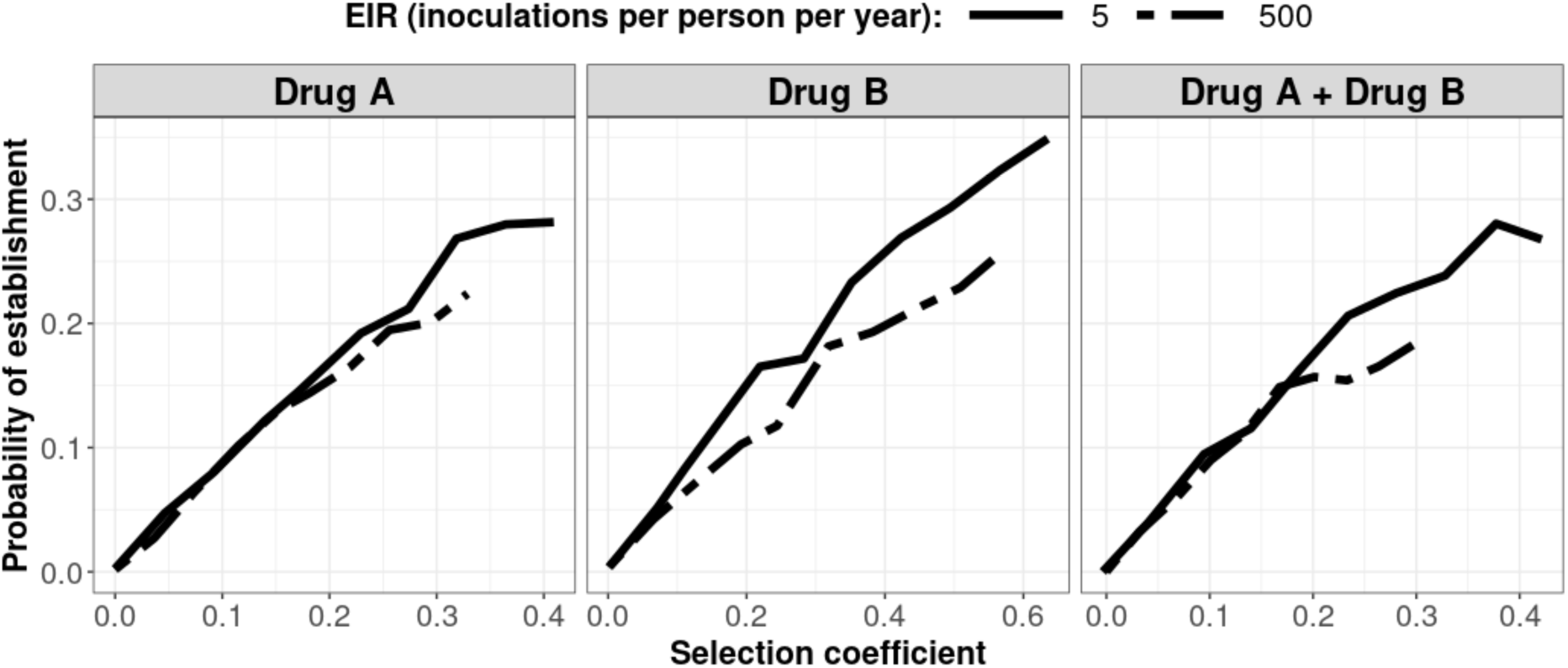
Predicted probability of establishment of mutations conferring drug resistance across transmission settings. Solid curves and dashed curves represent the relationship between the selection coefficient and the estimated probability of establishment of resistant parasites across settings that differ in transmission intensities (5 and 500 inoculations per person per year, respectively). The range of selection coefficients include higher values at a low EIR. For each setting, the level of access to treatment was specified as 80%, the population was assumed to be fully adherent to treatment (100%), and transmission was non-seasonal.

## Discussion

Understanding which disease, transmission, epidemiological, health system, and drug factors systemically drive the evolution of drug resistance is challenging. A full understanding requires vast observational data or clinical trials on a scale that is not possible or mathematical models that are sufficiently detailed to capture all these factors while remaining computationally feasible to simultaneously assess the impact of these factors. In response to this need, we updated a detailed individual-based model of malaria dynamics to include a full pharmacological (i.e. PK/PD) description of antimalarial treatments. We introduced a global sensitivity analysis approach based on emulators for computationally intensive models to systematically assess which factors jointly drive the evolution of drug-resistant parasites. As discussed below, our approach allowed us to understand the guiding principles of the evolution of drug resistance against ACTs and to explain the difference in trends observed in the Greater Mekong Subregion (GMS) and in malaria endemic Africa. Improving our understanding of the factors that lead to drug resistance establishment and spread allows us to identify strategies to mitigate these dynamics and guides initial considerations for developing more sustainable malaria treatment.

Our results support the belief that evolution of resistance to ACTs begins with the establishment and spread of parasites resistant to the partner drug and, once the protective effect of the partner drug is reduced, drug selection falls on the artemisinin component, and parasites then start to acquire resistance to artemisinin derivatives (e.g. [48, 59]). The fact that resistance to the partner drug appears before resistance to artemisinin derivatives was supported by two points elucidated in our study. First, resistance to the partner drug strongly depends on the period of low concentration of this drug during which only resistant parasites can multiply within the host (known as the selection window). As artemisinin derivatives are short-acting, they cannot prevent patients from being reinfected by parasites resistant to the partner drug during this selection window. Second, resistance to the partner drug was the most critical factor that enhanced establishment and spread of partial artemisinin resistance. Without resistance to the partner drug, parasites partially resistant to artemisinin could only spread at a low rate as the partner drug could still eliminate them, thereby removing their selective advantage. Our results are in line with recent molecular data which show that parasites resistant to partner drugs (piperaquine and mefloquine) were already present in the GMS before partial artemisinin resistance emerged and that the spread of resistance to artemisinin accelerated when it became linked to resistance to the partner drugs [60–62]. Thus, the presence of partner drug resistance has probably facilitated the spread of resistance to artemisinin in the GMS. In contrast, in Africa, to date, only a low degree of resistance to the most commonly used partner drugs (lumefantrine and amodiaquine) are present [2, 63], which has likely limited establishment of resistance to artemisinin derivatives. We additionally note that the evolution of drug resistance in the GMS may have been favoured by the low transmission intensity (annual EIR range approximate from less than 1 to 25 inoculations per person per year [64–66]) compare to Africa where the transmission intensity is overall higher (annual EIR range from less than 1 to more than 500 inoculations per person per year [67, 68]). Similar to previous studies [4, 25, 28, 34, 69], establishment of drug resistance in our model was more likely in low transmission settings due to the reduced level of within-host competition between genotypes, as well as population immunity.

Our results suggest that a key strategy to mitigate the evolution of partial artemisinin resistance is to ensure that the partner drug efficiently kills the partially resistant parasite. Therefore, to delay the establishment of artemisinin resistance in Africa and to mitigate the spread of partial artemisinin resistance in regions where it is already established, we should ensure that limited or no genotypes are resistant to the partner drug for first-line ACT. One approach to ensure this is to implement robust molecular surveillance of resistance markers and to specify more sustainable treatment policies, such as changing first-line ACTs upon detection of resistance or when the frequency of resistant parasites reach a threshold as recommended by the WHO [2]. Furthermore, consistent with our results, adherence should continue to be promoted, as lower treatment compliance can lead to treatment failure even in the absence of resistance to the partner drug [2, 70, 71].

Our results suggest that future antimalarial therapies should shorten the selection windows of long-acting partner drugs. We show that resistance to long-acting drugs (drug B) is the first step in the evolution of resistance to ACTs, and it depends mainly on the length of the selection window. We confirm that the selection window strongly depends on the drug half-life, also consistent with previous studies [4, 23, 27, 28, 49, 58]. Consequently, reducing the half-life of the partner drug in an ACT regimen could reduce the spread of resistance. However, unless selection windows are substantially minimised or completely eliminated, the evolution of resistance would not totally be prevented [58]. Thus, a more sustainable option for ACTs would be to use triple artemisinin-based combination therapies (TACTs). TACTs involve combining an artemisinin derivative with two long-acting drugs [72].

If or when TACTs are to be widely used, our results emphasize that the two long-acting drugs should have matching half-lives to ensure that parasites are not exposed to residual drug concentrations of only one of the two partner drugs (noting that this is simple in principle, but more difficult in practice [73]). In addition, the parasite population should be devoid of parasites resistant to either of the two long-acting drugs. If resistance to one partner drug already exists in the population, the second partner drug would not be protected, and mutations conferring resistance to this second drug would be selected. Thus, ideally, we should avoid combining previously used long-acting drugs (because resistance to these drugs may already be present), and rather favour two partner drugs not routinely in use, and preferably having a new mechanism of action. Therefore, selecting drugs to combine for TACTs requires balancing the need to mitigate resistance against the development time of new partner drugs. Note that the development of new partner drugs for TACTs may be challenging because combining three drugs is likely to increase the risk of toxicity and the treatment price, and future antimalarial medicines must remain tolerated by patients and affordable [72].

Another approach to delay the evolution of partial artemisinin resistance could focus on extending the period of action of artemisinin derivatives. In our monotherapy analysis on the spread of a genotype partially resistant to artemisinin, we found that the spread of partially resistant genotypes decreased when the drug was present in patients for a longer time, such as if it had a long half-life and there was proper treatment adherence. This result arises because partially resistant parasites are still affected by the drug [40, 42–44]. Thus, increasing their exposure to the drug leads to higher killing and reduced spread. Increasing the exposure to artemisinin derivatives can be achieved by using the artemisinin derivative having the longest half-life and, as highlighted in other studies [74–76], can be done by increasing the number of doses and days that patients receive treatment. However, it is worth noting that extending the dosage regimen will be efficient only with adequate adherence to treatment, which may be challenging to achieve in practice. Also, as artemisinin derivatives are co-administrated with at least one long-acting drug, increasing the number of doses of this combination therapy would require reducing the concentration of the partner drug to prevent the partner drug from reaching toxic concentrations.

The evolution of drug resistance is a three-step process consisting of mutation, establishment, and spread. Mutation rates in malaria can easily be measured, and spread, quantified by the selection coefficient, is also easy to measure. However, the probability of establishment and its relation to the selection coefficient constituted a significant knowledge gap. Standard population genetic models assume that the number of secondary infections follows a Poisson distribution [15, 77]. Under this assumption, for selection coefficients lower than 0.2 (according to an informal literature review in [16], most selection coefficient estimates for malaria drug resistance mutations from the field fall between 0.02 to 0.12), the probability of establishment is approximately equal to twice the selection coefficient [15, 77]. However, the number of secondary malaria infections more likely follows a negative binomial distribution due to the high heterogeneity of transmission, which may substantially reduce the probability of establishment (Box 2 of [15]). In this modelling study, we were uniquely able to quantify the link between selection coefficients and the probability of establishment of mutations. On average, we predicted that, for selection coefficients lower than 0.2, the probability of establishment was equal to 0.87-times the selection coefficient. Therefore, our findings suggest that the variation in the number of secondary infections of *P. falciparum* must be much greater than the Poisson distribution assumed by standard population genetics models, and this higher variation reduces the probability of establishment of emerging mutations.

As with all modelling studies, our approach has several limitations, primarily arising from constraints imposed by the model. First, our drug action model does not capture stage-specific killing effects, so we could not model parasites partially resistant to artemisinin being insensitive to the drug only during extended ring-stage [40, 42–44], although previous analyses suggested this would be captured by our variation in the maximum killing rate [78]. Nevertheless, if we modelled a reduction of the drug effect restricted to the ring-stage, we expect to obtain similar results. That is, a long half-life and high treatment adherence would increase the likelihood that the drug is present within patients during any stage other than the ring-stage, and thus the drug would kill more resistant parasites.

Second, our model did not capture the impact of artemisinin resistance on gametocytes. Previous studies have highlighted that artemisinin kills gametocytes, and patients infected with parasites partially resistant to artemisinin exhibit higher gametocyte densities than patients infected with sensitive parasites [79, 80]. We did not model the impact of artemisinin and resistance on gametocytes. This effect is likely to accelerate the spread of partial resistance. However, the relationship between the different factors reported in this study should be unchanged.

Third, our model, OpenMalaria, does not capture the recombination of *P. falciparum* parasites in mosquitoes. Currently, OpenMalaria does not support chromosomal recombination as it does not track the different genotypes in mosquitoes, and the genotype of new infections is based on the genotype frequency in humans. Previous models have shown that if multiple mutations are needed to confer drug resistance, recombination could slow the evolution of drug resistance by separating these mutations, especially in settings with a high rate of infection [69, 81]. We ignored the effect of recombination by assuming that only one mutation differs between the resistant and sensitive genotypes, which is valid for resistance to certain drugs, such as artemisinin [2, 3]. However, for other drugs, such as sulfadoxine-pyrimethamine, drug resistance is due to the accumulation of multiple mutations in two genes [82].

Lastly, to investigate the establishment of drug-resistant parasites, we modelled the emergence of mutations through importation. Consequently, our estimations represent the establishment of mutations imported into a population or mutations emerging in mosquitoes (assuming that the mosquito has only transmitted the mutated genotype and not the wild type genotype to the individual). A mutation emerging during the blood-stage within the human host may have a lower probability of establishment because sensitive parasites would be present in the host, leading to competition between them. It is still unclear whether mutations conferring drug resistance arise during the blood-stage (due to the high parasite numbers) or during the sexual stage in mosquitoes (because recombination generates many genetic variations). Nevertheless, the probabilities of establishment estimated in this study are consistent with the probabilities of establishment predicted by a previous study [15].

In summary, our results confirm that mutations conferring malaria drug resistance are more likely to establish in low transmission settings. Our results demonstrate that the establishment and spread of resistance to artemisinin derivatives have likely been facilitated by pre-existing resistance to partner drugs. Thus, it is essential to prioritise monitoring and to limit the spread of resistance to partner drugs in current or future ACT regimens. If resistance to the partner drug is confirmed, response strategies should prioritise monitoring molecular markers and treatment failure and switching to an ACT with an effective partner drug should be considered. In addition, our results show that drug properties play an essential role in the evolution of parasite drug resistance. Thus, the ongoing development of new antimalarial combinations should limit selection windows of partner drugs by matching half-lives, hopefully leading to longer lasting combination treatments against malaria. In the medium-term, for existing ACTs, it would be advantageous to increase the time of parasite exposure to the short-acting artemisinin derivate and/or to include a second long-acting partner drug with a matching half-life to the other long-acting partner drug (triple ACTs [72]) and for which limited or no parasite resistance exists in the target population.

## Methods

### Simulation model and the parameterisation of treatment profiles and resistant genotypes

#### (i) Overview of our OpenMalaria model

Our individual-based model, OpenMalaria, simulates the dynamics of *P. falciparum* in humans and links it to a periodically forced deterministic model of *P. falciparum* in mosquitoes [83–85]. The model structure and fitting are described in detail elsewhere [84, 85], including open-access code (https://github.com/SwissTPH/openmalaria) and documentation (https://github.com/SwissTPH/openmalaria/wiki), and a recently published manuscript provides a new calibration [86]. Here, we have summarised the main components of OpenMalaria and its latest developments in version 40.1, which enabled us to model the establishment and spread of drug-resistant parasites.

OpenMalaria is an ensemble of models in which mosquito and infection events, and parasite and human attributes are updated every five days. A demography model maintains a constant human population size and age structure across the simulation. Multiple parasite genotypes and their initial frequency can be defined in more recent model versions. For each infection, a mechanistic model simulates the parasite dynamics within the host and incorporates innate, variant, and acquired immunity [50]. The within-host model allows for concurrent infection of multiple parasite genotypes within the same host and captures indirect competition between genotypes based on host immunity, which regulates the overall parasite load. The user can specify a reduction of the within-host multiplication factors of each genotype to model a fitness cost associated with the mutation. The host’s parasite density determines the symptoms and mortality of patients and diagnostic test results. The occurrence and severity of patient symptoms depend on their pyrogenic threshold, which increases (until saturation) with recent parasite exposure and decays over time [87]. Severe episodes of malaria occur due to a high parasite density or due to co-morbidities [88]. Malaria mortality can be a consequence of a severe episode or an uncomplicated episode with co-morbidity [88, 89]. The model also takes into account neonatal deaths [88, 89]. Immunity to asexual parasites prevents severe cases by decreasing the parasite multiplication rate within the host. Individual immunity depends on the cumulative parasite and infection exposure frequency, as well as maternal immunity in their newborn children for several months [90].

The case management component of OpenMalaria describes the use of treatment for uncomplicated and severe cases and depends on access to health services and whether patients have previously been treated for the same episode [91]. The disease model includes explicit pharmacokinetic-pharmacodynamic (PK/PD) models that capture the process whereby drugs reduce the parasite multiplication rate in treated hosts [51, 54]. Pharmacodynamic parameters are parameterised individually for each genotype to allow different degrees of drug susceptibility to be modelled.

The entomological component of OpenMalaria simulates the mosquito vector feeding behaviours and tracks the infectious status of mosquitoes [83]. The periodicity of this model allows seasonal patterns of transmission to be captured. The probability that a feeding mosquito becomes infected depends on the parasite density within bitten individuals [92]. No recombination is modelled between the different genotypes in the mosquitoes. The number of newly infected hosts depends on the simulated entomological inoculation rate (EIR) of the vector model [83]. The genotype of new infections is based on the genotype frequencies in humans from the previous five time steps [92].

#### (ii) Parameterisation of the treatment profiles

This study investigated factors influencing the establishment and spread of parasites resistant to three different treatment profiles.

The first treatment profile modelled was a short-acting drug administered as monotherapy, referred to as drug A. Drug A has a short half-life and a high killing efficacy, simulating artemisinin derivatives (Figure 1A and 1B). We modelled the pharmacokinetics of drug A using a one-compartment model, which is considered sufficient when modelling short-acting antimalarials [52, 54]. We varied key PK/PD parameters (half-life, EC50, Emax) in the global sensitivity analysis to assess their influence on the rate of spread of resistance. The EC50 ranged from 0.0016 to 0.009 mg/l to include the EC50 of artemether, artesunate, and dihydroartemisinin [52, 54]. The half-life parameter ranges represented the values for artemether, artesunate, and dihydroartemisinin used by [52, 54] (Table 1). Note that in [52], the Emax of all short-acting drugs was equal to 27.6 per day. However, we varied the killing rate and included higher values to investigate its effects on the rate of spread (Table 1). To ensure that drug A killed the sensitive parasites efficiently for any combination of parameters, we extended the treatment course from a daily drug dose for three days to a daily drug dose for six days. Moreover, we parameterised the dosage and constant parameter values to that for dihydroartemisinin (Table S1), as it is the artemisinin derivate with the shortest elimination half-life and highest EC50 [52, 54]. By doing so, we also ensured that drug A had the typical profile of an artemisinin derivative.

The second treatment profile modelled was a long-acting drug administered as monotherapy, referred to as drug B. Drug B had a long half-life and a lower Emax than drug A (Figure 1A and 1B), typical of partner drugs used for ACTs. We modelled the PK of drug B with a two-compartment model, which is more typical of the clinical PK of partner drugs [51]. As for drug A, key PK/PD parameters (half-life, EC50, Emax, and dosage) were varied in the global sensitivity analysis. The EC50 ranged from 0.01 to 0.03 mg/l to include the EC50 of mefloquine, piperaquine and lumefantrine used by [52, 54]. The half-life range corresponded to the value reported for mefloquine, piperaquine, lumefantrine in [93–97] (Table 1). We increased the Emax range from 3.45 per day (as reported in [54]) to 5.00 per day to investigate the effect on the rate of spread (Table 1). We also assessed the impact of Cmax on the rate of spread for drug B because the Cmax varies between ACTs partner drugs and has a strong influence on the post-treatment killing effect of drug B [73]. We varied drug dosage from 30 to 40 mg/kg to examine the influence of variation of Cmax on the spread rate for drug B. The lower limit of 30 mg/kg was fixed to ensure that drug B killed the sensitive genotype efficiently for any parameter combination. The treatment course involved a daily drug dose for three consecutive days. To ensure that drug B had the profile of typical partner drugs, the values of the constant parameters were parameterised to the values of piperaquine reported in [54, 94] (Table S2).

The last treatment profile was a combination of drugs A and B, simulating ACT. We tracked the concentration of each drug independently. We used the same models, parameter values and ranges for the two drugs as when both drugs were used as monotherapy. However, the treatment course involved a daily dose of both drugs for three days, as recommended by the WHO for most ACTs [56]. In OpenMalaria, the killing effects of the two drugs were calculated independently and acted simultaneously on the parasites.

#### (iii) Parameterisation of the drug-resistant genotypes

For each simulation, we tracked two genotypes, one drug-resistant and one drug-sensitive. We investigated the spread of resistant parasites with different degrees of resistance (Table 1). We modelled the phenotype of drug resistance and the degree of resistance differently for each drug profile.

Previous studies have shown that parasites partially resistant to artemisinin exhibit an extended ring-stage during which they are not sensitive to artemisinin (even at high drug concentrations) but remain sensitive to the drug during other stages of the blood replication cycle [40–44]. OpenMalaria does not model the specific drug-killing effect for the different steps of the blood-stage. As in [98, 99], we assumed that parasites resistant to drug A had a reduced Emax compared with sensitive ones (Figure 1B). This assumption captured the fact that, overall, drug A killed fewer resistant parasites than sensitive ones at any drug concentration because they are not sensitive to artemisinin during the ring-stage and that this stage-specific effect is best incorporated into PK/PD modelling by variation in Emax [78].

Previous studies reported that parasites resistant to long-acting drugs typically have an increased EC50 [45–47]. Thus, as in other models, we defined parasites resistant to drug B to have a higher EC50 than the sensitive ones (Figure 1B) [52, 54]. With an increased EC50, the resistant parasites were less susceptible to the drug at low drug concentrations. Thus, these resistant genotypes were more likely to survive drug treatment and are more likely to successfully infect new hosts with higher residual drug concentrations [58].

Considering drugs A and B in combination, the resistant genotype was resistant to drug A. But in the global sensitivity analysis, both the sensitive and resistant genotypes could have some degree of resistance to drug B. The decreased susceptibility to drug B was the same for both sensitive and resistant genotypes, meaning that we assumed the two genotypes differed only in one mutation, which conferred resistance to drug A. This assumption allowed us to ignore the effect of recombination in the mosquitoes. In effect, this assumed that the allele defining the level of resistance to drug B was fixed in the population.

### Approach to identify the key drivers of the spread of drug-resistant parasites

Through global sensitivity analyses, we quantified how the factors in Table 1 influenced the spread of drug-resistant parasites for each treatment profile. First, we estimated the effect of each factor in a non-seasonal setting with a population fully adherent to treatment. Based on these results, we identified specific settings for further analysis. We performed additional constrained sensitivity analyses to investigate the impact of varying drug properties and fitness costs in a fixed set of settings (i.e. in low and high transmission settings, with low and high treatment levels of monotherapy or combination therapy) and with a fixed degree of resistance. In this secondary analysis, we also investigated the effect of drivers in seasonal transmission settings (Figure S10) and where populations adhere to either 100% or 67% of treatment doses.

Due to the computational requirements for a large number of simulations of OpenMalaria, and the number of factors investigated, it was not feasible to simulate either a full-factorial set of simulations to perform a multi-way sensitivity analysis, or to perform a global sensitivity analysis. Therefore, we trained a Heteroskedastic Gaussian Process (HGP) [100] on a set of OpenMalaria simulations and performed global sensitivity analyses using this emulator (Figure 1C), adapting a similar approach to [101] and [86]. Our approach involved: (i) randomly sampling combinations of parameters; (ii) simulating and estimating the rate of spread of the resistant genotype for each parameter combination in OpenMalaria; (iii) training an HGP to learn the relationship between the input (for the different drivers) and output (the rate of spread) with iterative improvements to fitting through adaptive sampling, and (iv) performing a global sensitivity analysis based on the Sobol variance decomposition [70]. Each step of the workflow is detailed below.

#### (i) Randomly sample combinations of parameters

We randomly sampled 250 different parameter combinations from the parameter space shown in Table 1 using a Latin Hypercube Sampling (LHS) algorithm [57]. The parameter ranges were defined as follows. We defined the ranges for the properties of drug A and drug B to include the typical parameter values of artemisinin derivatives and a long-acting partner drug, respectively [52, 54, 93–97]. The range of the degree of resistance captured the spread of drug-resistant parasites, which vary from fully sensitive to having almost no drug sensitivity. The fitness costs were extracted from studies investigating the decline of chloroquine-resistant parasites after the drug pressure was removed [102, 103]. The variation in annual EIR captured settings with low transmission to those with high transmission. The range of access to treatment captured settings with low to high 14-days effective coverage. The values of the diagnostic detection limit captured the sensitivity of typical diagnostics used for malaria (rapid diagnostic test, microscopy, and polymerase chain reaction (PCR)) [104, 105].

#### (ii) Simulate and estimate the rate of spread of the drug-resistant genotype

We quantified the rate of spread through the selection coefficient, a measure widely used in population genetics to assess the strength of selection on a genotype [16]. The selection coefficient is the rate at which the logit of the resistant genotype frequency increases each parasite generation and should be linear throughout the spread [16]. Population genetics theory often assumes an infinite population size to remove stochastic fluctuation of the allele frequency, also called genetic drift [16]. However, in our model the parasite population size is finite, so stochastic fluctuations of the genotype frequency are present. Thus, we should avoid estimating the selection coefficient when there is a low frequency of the resistant genotype (from a small human population size, a low EIR, and a small initial frequency of the resistant genotype) because the resistant genotype may become extinct due to the stochastic fluctuation. In addition, the effects of genetic drift that occurs when a genotype is present at a low frequency may cause non-linearity during resistance spread which may obscure the estimation of the selection coefficient [16].

Following the approach described in [16], we assumed an initial percentage of infected humans carrying the resistant genotype of 50%. A high initial percentage minimises the impact of random fluctuation on our estimation, and the subsequent risk of extinction, without affecting our estimate because the selection coefficient was not frequency-dependent (Figure S11). We simulated the spread of resistant parasites in a human population of 100,000 individuals with an age structure of a typical African country [106]. We ran each parameter combination on five stochastic realisations. The simulation started with a burn-in period of 100 years to reach the expected level of immunity in the population and an additional 30 years to reach EIR equilibrium (Figure S12). Both genotypes were sensitive to the drug during this period, so the percentage of infected humans carrying the resistant genotype remained stable. After the burn-in period, we introduced the fitness cost and the drug for which the resistant genotype had reduced sensitivity. We then estimated the selection coefficient, *s*, as,

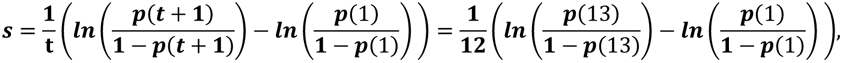

where *p*(*t*) is the relative frequency of the resistant genotype in inoculations, *t* is the number of parasite generations after introducing the new drug at *t* = 0. We assumed that a parasite generation is two months (60 days) as in [16]. We started the regression at one parasite generation after introducing the new drug (at 60 days). We stopped the regression 12 generations later, at 720 days, because, as shown in [16], it was computationally convenient and returned stable selection coefficient estimates. The regression was stopped sooner if the relative frequency of inoculations carrying the resistant genotype was higher than 90% or lower than 30% to prevent tracking a small number of a single genotype for which genetic drift is strong. In seasonal settings, the rate of spread of the resistant genotype varied throughout the year. Consequently, we estimated the selection coefficient using a moving average of the relative frequency of the resistant genotype in inoculations (Figure S13). This method prevented biasing the selection coefficient according to the period included in the regression.

Once the selection coefficient was estimated, it could be converted to the number of parasite generations needed for the relative frequency of the resistant genotype in inoculations to increase from *p(1)* to *p(t)*,

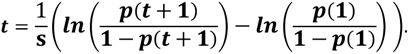

We could then convert the number of parasite generations to time in years, a more relevant public health measure than the selection coefficient itself.

#### (iii) Train the emulator and improve its accuracy

We randomly split our data into a training dataset containing 80% of simulations and a test dataset containing 20% of simulations. We trained the Heteroskedastic Gaussian Process (HGP) on the training dataset using the function mleHetGP from the R package ‘hetGP’ [100]. To assess the accuracy of the emulator, we predicted the selection coefficient for the test dataset with the emulator and compared these predictions with the expected selection coefficient estimated using OpenMalaria. We iteratively improved the accuracy of our emulator through adaptive sampling. Adaptive sampling involved resampling 100 parameter combinations in the parameter space where we were less confident (higher variation) in the HGP prediction and repeating the entire process until the emulator had a satisfactory level of accuracy. The satisfactory level of accuracy was defined based on the correlation coefficient and the root means squared error of the predicted selection coefficient and expected selection coefficient (Figure S14-20).

#### (iv) Global sensitivity analysis

Using the emulator, we undertook global sensitivity analyses using Sobol’s method [107]. This method attributed fractions of the selection coefficient variance to each input [107]. We performed the global sensitivity analysis using the function soboljansen from the R package ‘sensitivity’ [108], and two random datasets with a sample size of 100,000, with 150,000 bootstrap replicates. With this function, we estimated first-order and total Sobol’ indices simultaneously. The first-order indices represent contributions of each parameter’s main effect to the model output variance. The total effect represents the contribution of each parameter to the model output variance considering their interactions with other factors. We report only the first-order indices in the Results section because we did not observe many interactions between these factors. Some parameters supported the spread of resistance (increased the selection coefficient), whilst others hindered the spread (decreased the selection coefficient). To visualise the direction of the effect of each parameter, we calculated the 25th, 50th, and 75th quantiles of the predicted selection coefficient over the corresponding parameter ranges.

### Establishment of drug resistance

As explained in the Introduction, the establishment of resistant mutations is a stochastic process that depends on the selection coefficient of the mutation and the heterogeneity of parasites reproductive success in the setting, which in turn depends on the transmission level and the health system strength [13, 15–18]. Estimating the probability of establishment requires running many stochastic realisations due to the stochasticity of this step. To be more computationally efficient, we assessed the probability of establishment of a subset of 10 resistant genotypes with a known selection coefficient per setting and treatment profile. Based on the observed relationships between the selection coefficient and the probability of establishment for each treatment profile and setting, we could then extrapolate the probability of establishment of any mutations having a known selection coefficient.

To estimate the probability of establishment, we modelled the emergence of resistant mutations in a fully susceptible population. We used the approach described in [16] in which resistant infections were imported into the population at a low rate. In OpenMalaria, imported infections have the same frequencies of genotypes as in initialisation, thus we cannot import only resistant infections. Therefore, to import resistant infections in a population infected only by sensitive parasites, we followed the step described below (Figure S21). We first defined a 50% relative frequency of resistant parasites in infected humans. The simulation started with a burn-in phase of 100 years, during which both genotypes were sensitive to treatment. This meant that the relative frequency of the resistant parasites was stable (at 50%). In the second phase, we introduced a drug to which resistant parasites were hypersensitive (the drug EC50 was 100-times lower in the resistant genotype than the sensitive one). The second phase ran for 100 years, and once complete, the parasite population was fully susceptible. In the third phase, we imported new infections at a rate low enough to ensure that the previously imported mutation either established or went extinct before a new resistant mutation was imported (Supplementary file 1: section 5.1). The third phase ran until one mutation established (over 50% of infected humans carried the resistant genotype).

The probability of establishment, *P_e_*, can be estimated based on the average number of mutations that are imported until one mutation establishes, *N_e_*, as follows (the probability of a successful event can be estimated as one divided by the mean number of independent trials required to achieve the first success [109]),

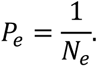

We simulated 300 stochastic realisations, *R*, and estimated *P_e_*, as,

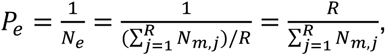

where *N_m,j_* is the number of imported mutations until one mutation established in run *j.* Re-arranging the formula shows that *Pe* is equal to the number of mutations established in all stochastic realisations (this number is equal to *R* as only one mutation established per stochastic realisation) divided by the total number of mutations imported into all stochastic realisations (mutations that became extinct and established). Note that in each stochastic realisation, we estimated *N_m_*, as,

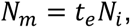

where *t_e_* is defined as the last time that the number of infections with a resistant genotype was equal to zero, i.e. the time (in years) until the arrival of the first mutation that successfully establishes. *N_i_* is the number of imported resistant infections per year. Note that OpenMalaria specifies the number of imported infections, *V*, in numbers of imported infections per 1,000 people per year, and half of the imported mutations were sensitive. Thus, the number of imported resistant infections that occurred until one established can be estimated as,

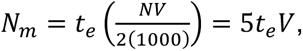

where *N* is the human population size. We set the population size to 10,000 to be computationally feasible (as since we were not measuring the selection coefficient, there was no need to minimise the influence of stochastic processes).

## Data Availability

We did not use individual participant-level data. Parameters values used in the model were informed from the literature as referred to in the main text or the supplementary file. The source code for OpenMalaria was developed using the C++ language and is available at https://github.com/SwissTPH/openmalaria. The analysis script was developed using the R software. All generated data and the code used for data analysis and visualization will be made available on request.

## Acknowledgements

We sincerely acknowledge all members of the Disease Modelling Unit of the Swiss Tropical and Public Health Institute and Dr Raman Sharma from Liverpool School of Tropical Medicine for their inputs. Simulations were performed on the scientific computing core facility, sciCORE, at the University of Basel (http://scicore.unibas.ch/).

## Author contributions

MAP Conceptualization. TM, MG, AJS, IMH, MAP Methodology; TM, MG, AJS Software; TM Validation; TM Formal analysis; TM, TL, IMH, and MAP Investigation; TM Data curation; TM, TL and MAP Writing – original draft preparation; TM, TL, MG, AJS, SLK, IMH, and MAP Writing – review & editing; TM Visualization; MAP Supervision; MAP Project administration; MAP Funding acquisition

## Supplementary file

## 1. Supplementary results

### 1.1 Supplementary figures that illustrate the results

**Figure S1.**
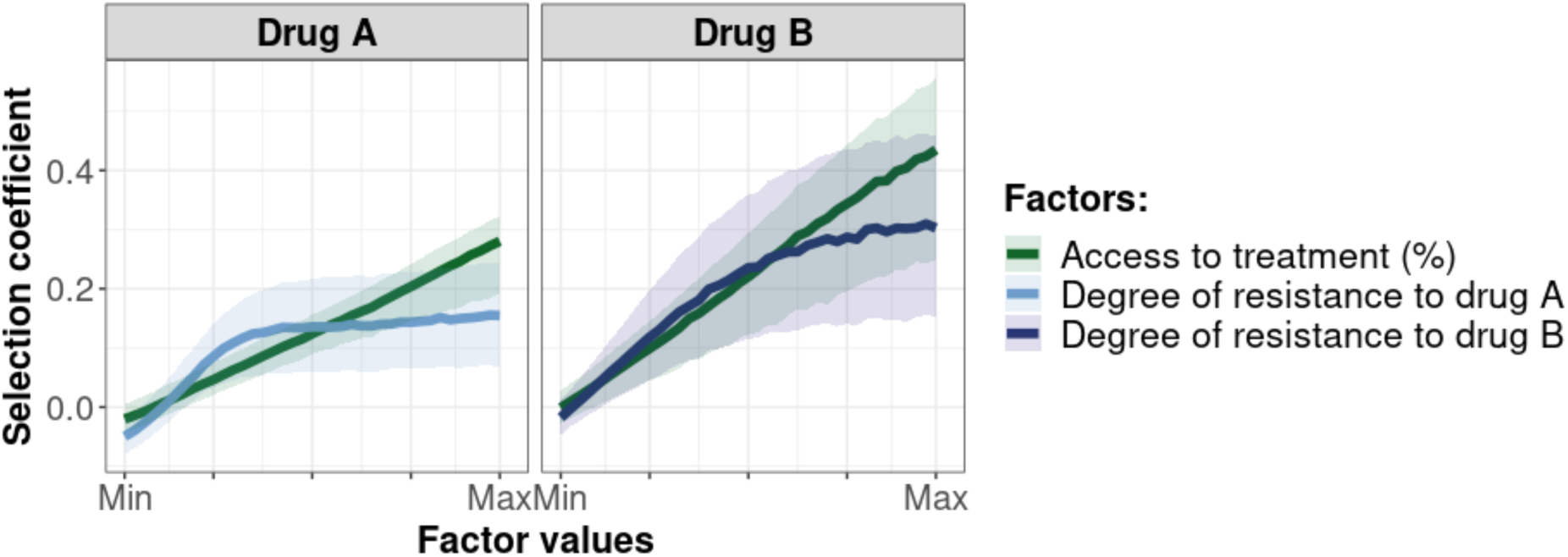
Influence of the access to treatment and degree of resistance on the selection coefficients of a genotype resistant to drug A or drug B used in monotherapy. Lines represent medians and shaded areas represent interquartile ranges of the selection coefficients estimated during the global sensitivity analysis over the parameter range for levels of access to treatment (10% to 80%), the degree of resistance to drug A (1- to 50-fold decrease in Emax), and the degree of resistance to drug B (1- and 20-fold increase in EC50).

**Figure S2.**
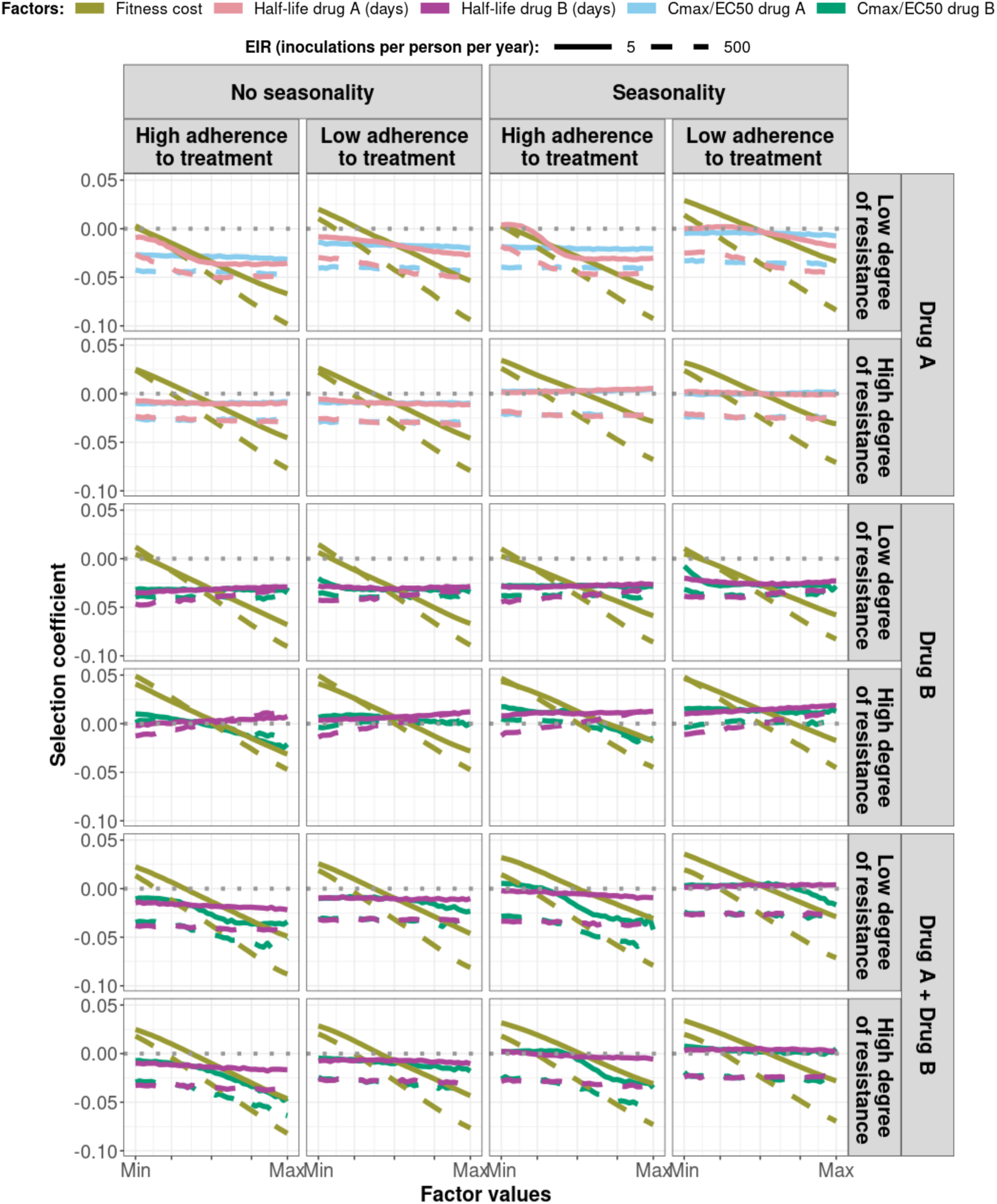
Magnitude and direction of effect of drug properties and fitness cost on predicted selection coefficients for low and high levels of transmission, degree of drug resistance, treatment adherence, in seasonal or perennial settings with monotherapy or combination treatment. The solid and dashed lines represent the median selection coefficients over the parameter ranges estimated in each setting that had low access to treatment (10%) and an *entomological inoculation rate (*EIR) of 5 (solid lines) or 500 (dashed lines) inoculations per person per year. Settings varied in their seasonality pattern and level of adherence to treatment *(low=67% and high=100%).* For each treatment profile, we show results for parasites with two different degrees of resistance; degree of resistance of 7 (low) and 18 (high) to drug A (Emax shift), 2.5 (low) and 10 (high) to drug B (EC50 shift), and with combination of drugs A and B, 7 (low) and 18 (high) to drug A and 10 to drug B. The parameter ranges were the following: fitness cost (1, 1.1); drug A half-life (0.035, 0.175) days; drug B half-life (6, 22) days; Cmax/EC50 ratio of drug A (55, 312); Cmax/EC50 ratio of drug B at a high level of adherence to treatment (5.4, 21.7); and at a low level of adherence (4.0, 16.2).

**Figure S3.**
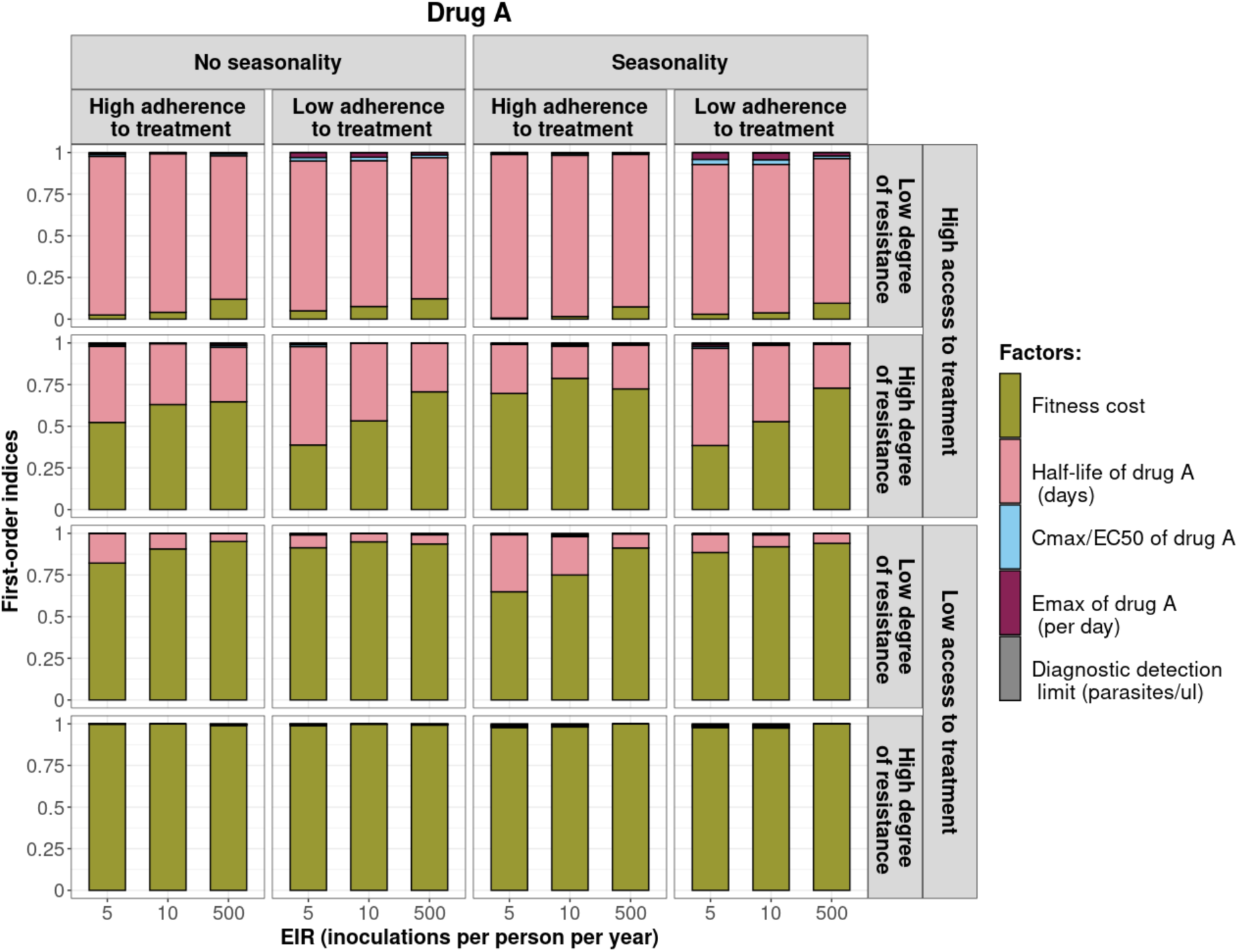
First-order indices describing level of importance of each factor from the constrained sensitivity analysis of the spread of a genotype resistant to drug A used in monotherapy. The first-order indices were assessed for parasites that had different degrees of resistance to drug A (low=7 and high=18 fold decrease in Emax) in settings that differ in their levels of access to treatment (high=10% and low=80%), levels of transmission (5, 10, and 500 inoculations per person per year), transmission patterns (no seasonality and seasonality), and levels of adherence to treatment (low=67%, and high=100%). The explored parameter ranges were the following: the fitness cost (1, 1.1); the half-life of drug A (0.035, 0.175) days; the ratio Cmax/EC50 for drug A (55, 312); the Emax of drug A (27.5, 31.0) per day; and the diagnostic detection limit (2, 50) parasites/microliter.

**Figure S4.**
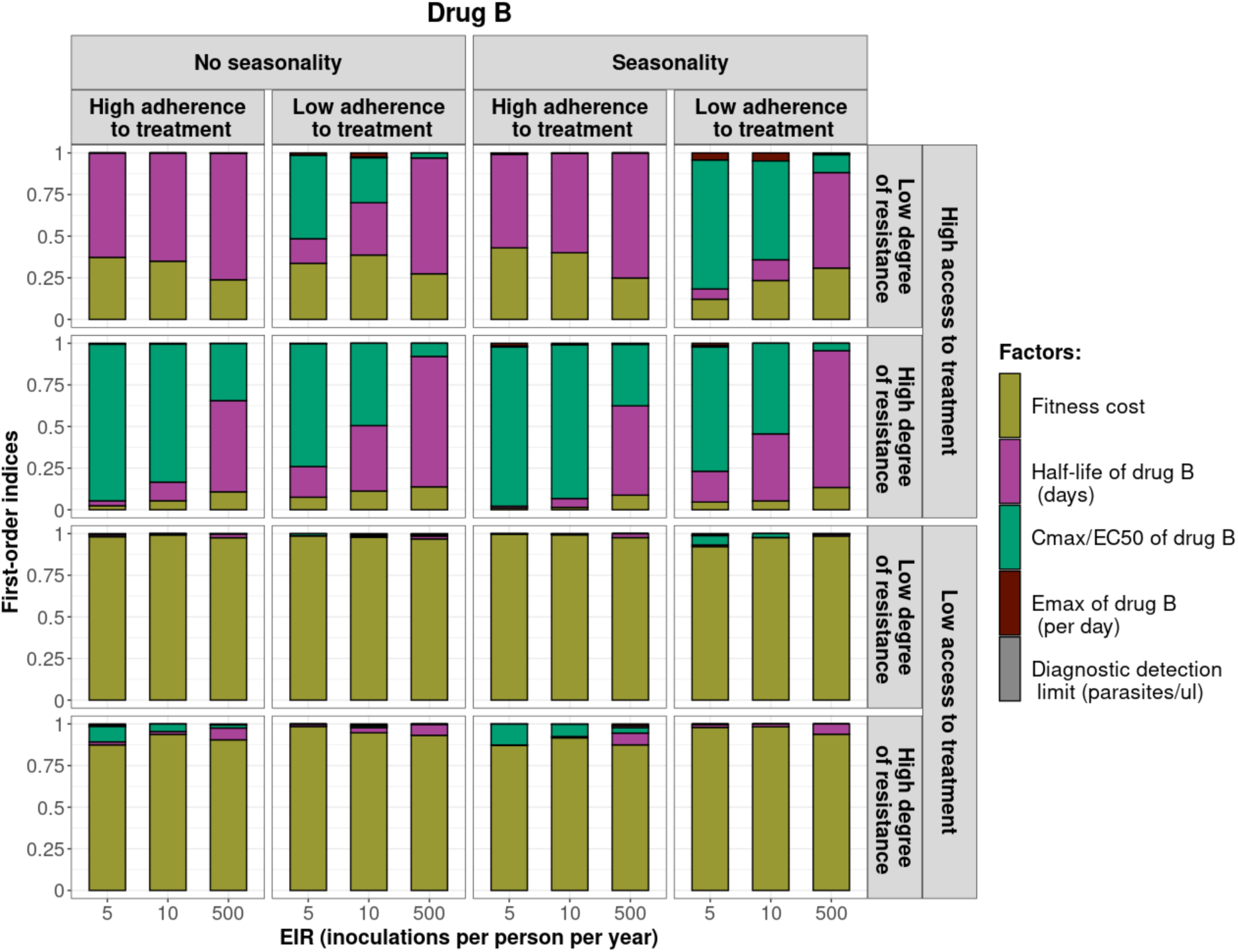
First-order indices of each factor from the constrained sensitivity analysis of the spread of a genotype resistant to drug B used in monotherapy. The first-order indices were assessed for parasites that had different degrees of resistance to drug B (low=2.5 and high=10 fold increase in EC50) in settings that differ in their levels of access to treatment (low=10 %, and high=80%), levels of transmission (5, 10, and 500 inoculations per person per year), transmission patterns (no seasonality and seasonality), and levels of adherence to treatment (low=67%, and high=100%). The explored parameter ranges were the following: the fitness cost (1, 1.1); the half-life of drug B (6, 22) days; the ratio Cmax/EC50 for drug B at a high level of adherence to treatment (5.4, 21.7) and at a low level of adherence to treatment (4.0, 16.2); the Emax of drug B (3.45, 5.00) per day; and the diagnostic detection limit (2, 50) parasites/microliter.

**Figure S5.**
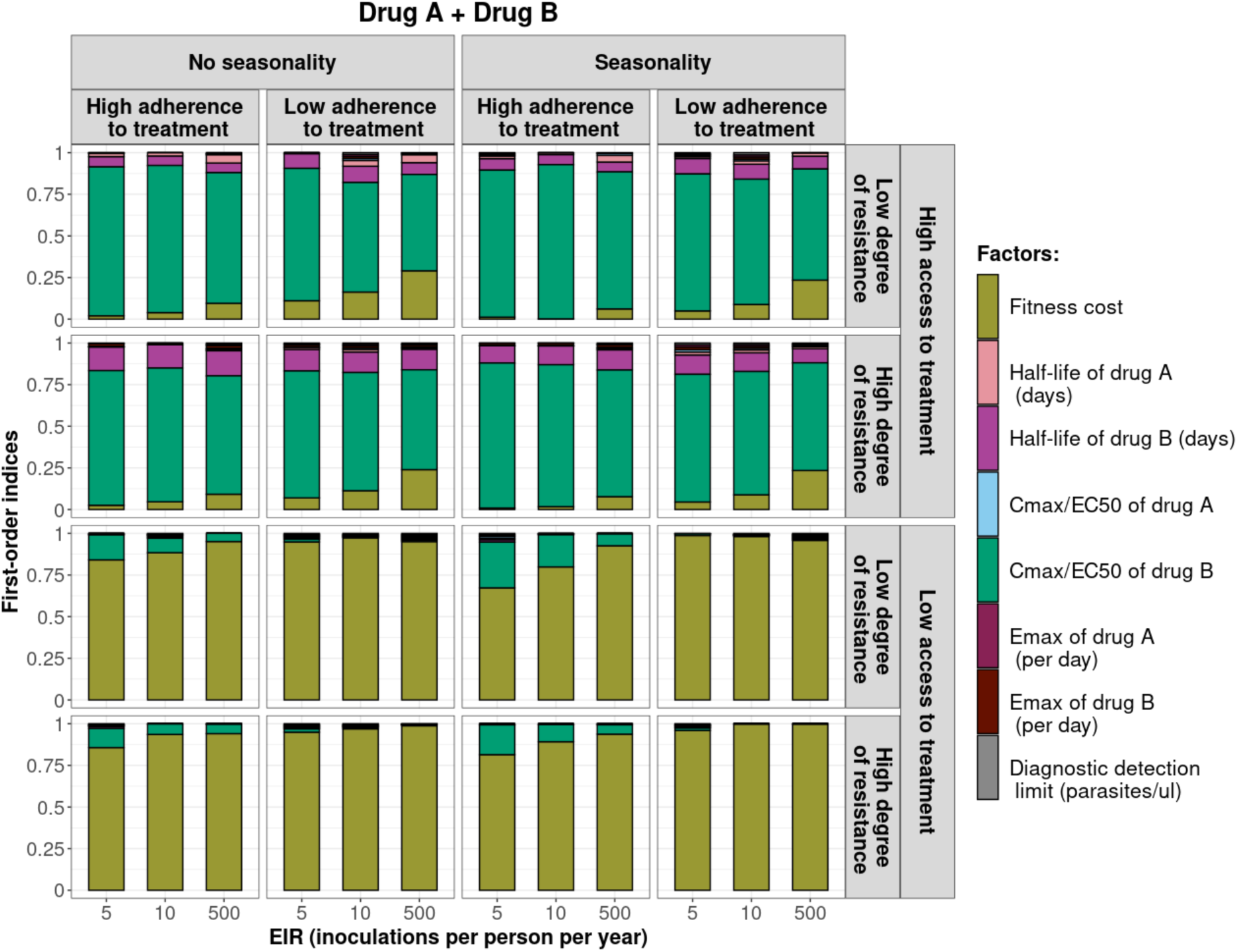
First-order indices of each factor from the constrained sensitivity analysis of the spread of a genotype resistant to drug A when drug A and drug B are used in combination. The first-order indices were assessed for parasites that had different degrees of resistance to drug A (low=7 and high=18 fold decrease in Emax) in settings that differ in their levels of access to treatment (low=10 %, and high=80%), levels of transmission (5, 10, and 500 inoculations per person per year), transmission patterns (no seasonality and seasonality), and levels of adherence to treatment (low=67%, and high=100%). The explored parameter ranges were the following: the fitness cost (1, 1.1); the half-life of drug A (0.035, 0.175) days; the half-life of drug B (6, 22) days; the ratio Cmax/EC50 for drug A (55, 312); the ratio Cmax/EC50 for drug B at a high level of adherence to treatment (5.4, 21.7) and at a low level of adherence to treatment (4.0, 16.2); the Emax of drug A (27.5, 31.0) per day; the Emax of drug B (3.45, 5) per day; and the diagnostic detection limit (2, 50) parasites/microliter.

**Figure S6.**
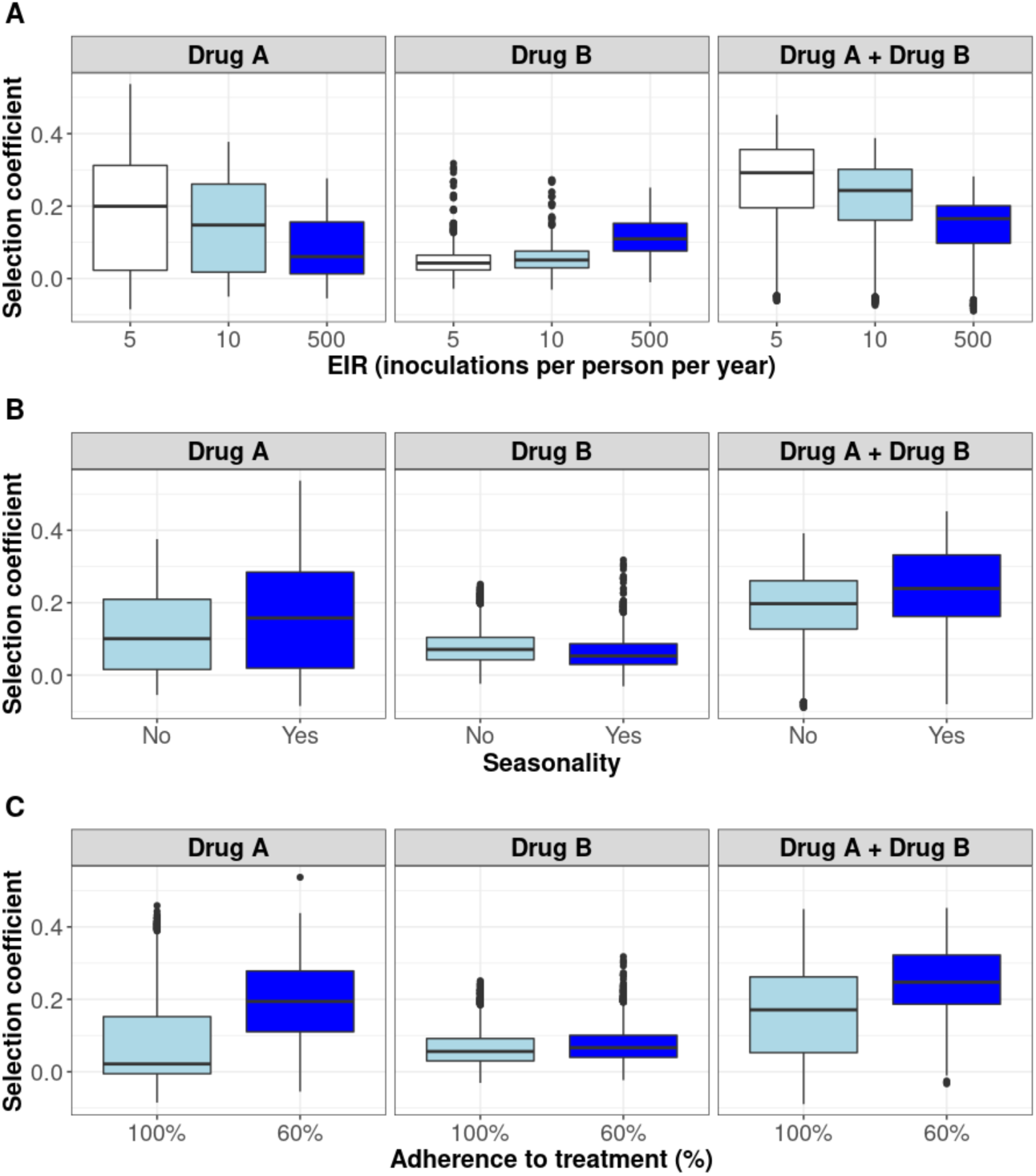
Distribution of selection coefficient across settings with high access to treatment. The selection coefficients were estimated for each treatment profile during the constrained sensitivity analysis of the spread of a resistant genotype having a low degree of resistance (equal to 7 for drug A (Emax shift), and to 2.5 for drug B (EC50 shift)), in settings with a high access to treatment (80%). The distributions are stratified by (**A**) the intensity of transmission (**B**) the seasonality pattern, and (**C**) the level of adherence to treatment in the settings.

**Figure S7.**
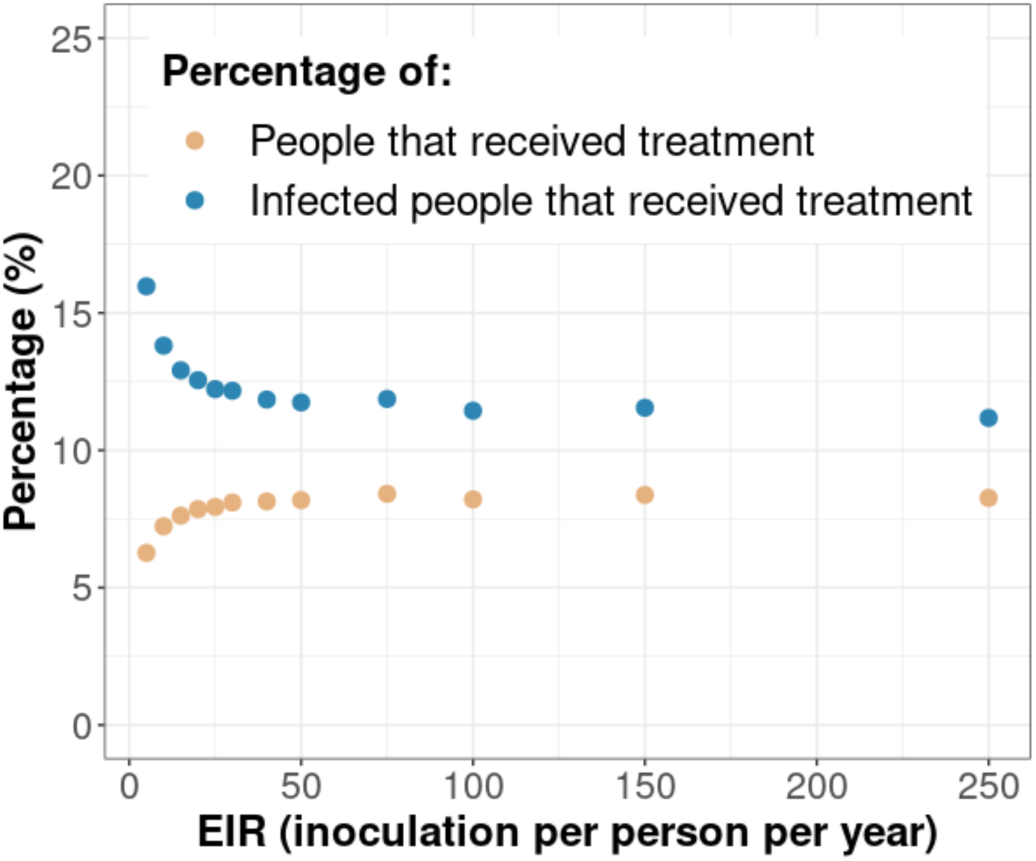
Treatment usage. The figure highlights the relationship between the transmission intensity (EIR) and the percentage of people that received treatment during a month (orange dots) and the percentage of infected people that received treatment during a month (blue dots). In this illustration, the level of access to treatment was equal to 80%, and the transmission was perennial.

**Figure S8.**
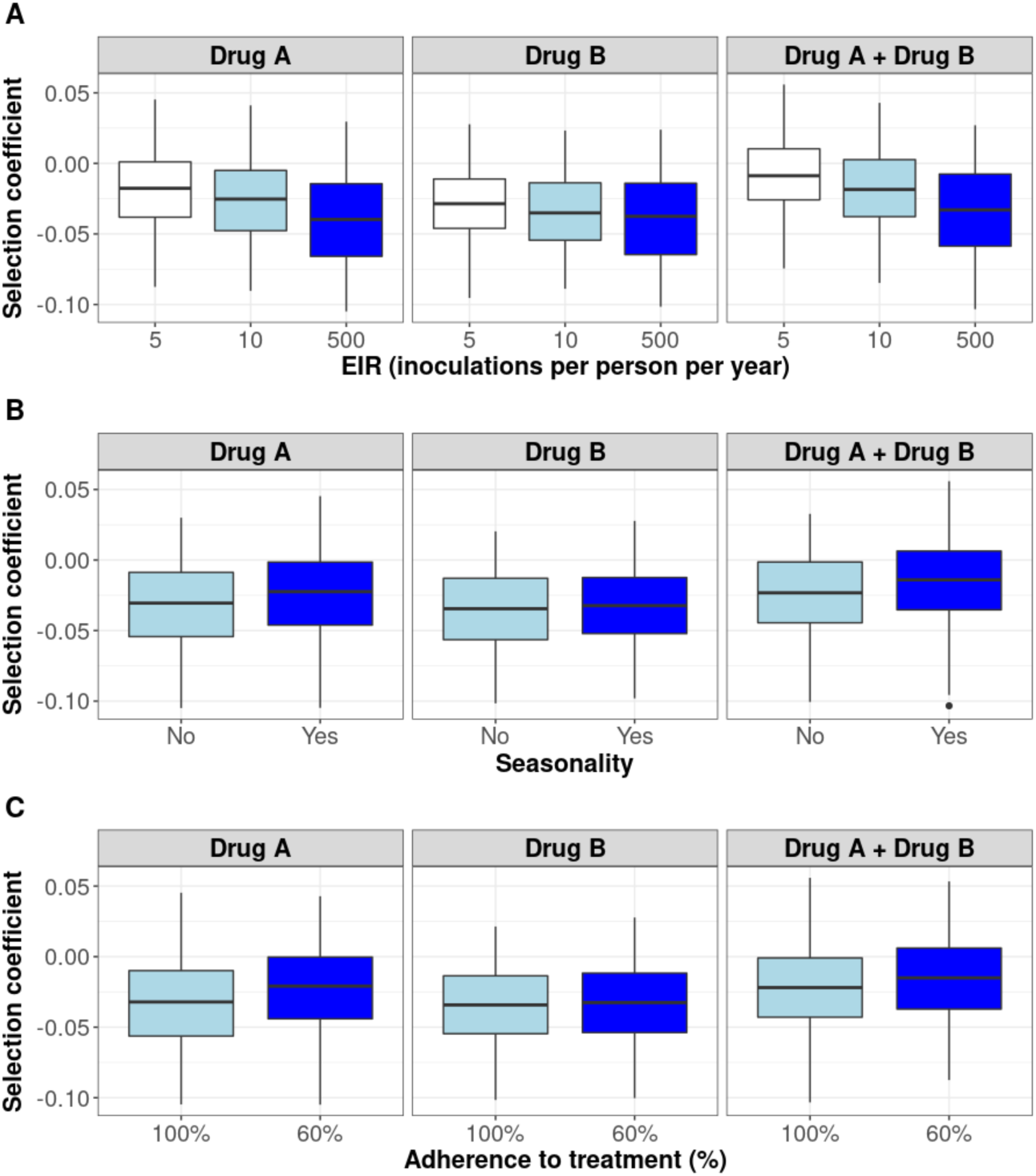
Distribution of selection coefficient across settings with a low access to treatment. The selection coefficients were estimated for each treatment profile during the constrained sensitivity analysis of the spread of a resistant genotype having a low degree of resistance (equal to 7 for drug A (Emax shift) and to 2.5 for drug B (EC50 shift)), in settings with a low access to treatment (10%). The distributions are stratified by (A) the intensity of transmission, (B) the seasonality pattern, and (C) the level of adherence to treatment in the settings.

### 1.2 The benefit of combination therapy

We illustrated the benefit of combination therapy by assessing how the degree of resistance to drug B influenced (i) the time taken for mutations conferring different degrees of resistance to drug A to spread from 1% to 25% of inoculations carrying the resistant genotype, *T25*, (Figure 5, first y-axis) and (ii) their probability of establishment (Figure 5, second y-axis). Both the *T25* and the probabilities of establishment were estimated based on selection coefficients predicted using the fitted emulators. To illustrate the impact of the transmission intensity on the two measurements, we predicted their values in low and high transmission levels. Note that, as discussed in the Results section, the relation between the selection coefficient and the probability of establishment changes slightly with the transmission level (Figure 4 of main text). In our example, drug A had the drug profile of dihydroartemisinin and drug B of piperaquine. We set the level of access to treatment to 100%, assumed no fitness cost, the transmission was perennial, and the population adhered to treatment fully.

In a low transmission setting, in a parasite population fully susceptible to drug B, parasites resistant to drug A had a low probability of establishment and required many years to spread from 1% to 25% of inoculations carrying the resistant genotype. For example, a mutation with a low (3.5-fold decrease in Emax) or high (13.5-fold decrease in Emax) degree of resistance to drug A had a probability of 1/1000 or 1/100, respectively, to establish in the population and required more than 39 years or over 18 years, respectively, to spread from 1% to 25% of inoculations carrying the resistant genotype (Figure 5). The probability of establishment and *T25* decreased tremendously with increased degrees of resistance of both genotypes to drug B (Figure 5). When the parasite population had a high degree of resistance to drug B (degree of resistance of 13.5), the probability of establishment increased to more than 1/10 and the *T25* was reduced to approximately three years, independent of the degree of resistance to drug A (Figure 5). These results confirm that resistance to partner drugs facilitates the establishment and spread of partial artemisinin resistance.

In high transmission settings, higher degrees of resistance to drug B also accelerated the establishment and spread of parasites resistant to drug A (Figure 5). However, the probability of establishment and the rate of spread were consistently lower in high transmission settings compared with low transmission settings (Figure 5). In addition, for a specific T50, the probability of establishment was slightly lower than in the low transmission setting due to the slight change in the relation between selection coefficients and probabilities of establishment with the EIR (see Results). These results agree with our observations that higher levels of within-host competition and immunity minimise the establishment and spread of resistance to artemisinin in high transmission settings.

**Figure S9.**
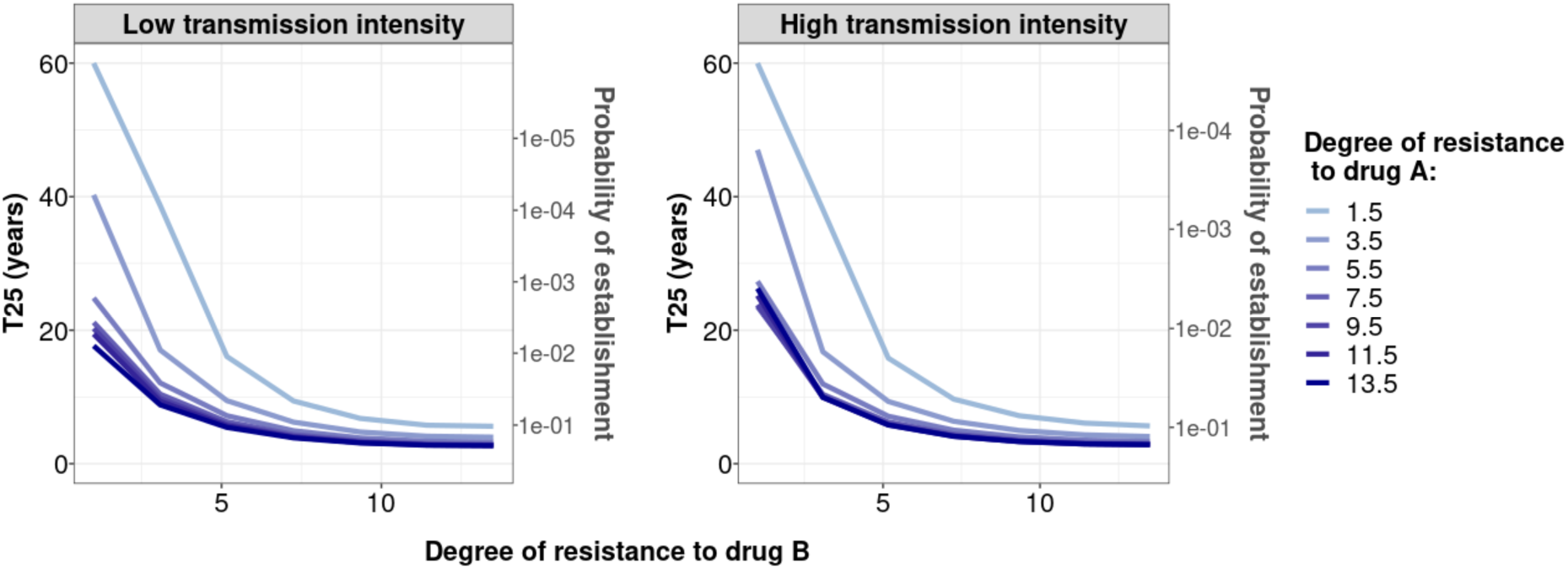
Illustration of the benefit of combination therapy on the evolution of drug resistance as time to 25% relative frequency of resistant genotypes. We estimated the probability of establishment and the time needed for parasites resistant to drug A to spread from 1% to 25% of inoculations carrying the resistant genotype, *T25,* for multiple degrees of resistance of the resistant genotype to drug A (Emax shift) and multiple degrees of resistance of both genotypes to drug B (EC50 shift). We assumed Drug A has a similar drug profile of dihydroartemisinin and drug B of piperaquine. We assumed a level of access to treatment of 100%. The population fully adhered to treatment. The resistant parasites had no fitness cost. The transmission intensity was equal to 5 (low transmission intensity) or 500 (high transmission intensity) inoculations per person per year (reflected low to very high transmission). The transmission was perennial.

## 2. Details on the parameterisation of OpenMalaria

**Table S1.** Pharmacokinetics (PK) and pharmacodynamics (PD) parameter values for drug A that were kept constant throughout the sensitivity analyses.

| Component | Parameter | Value | Reference |
| --- | --- | --- | --- |
| PK | Volume distribution (l/kg) | 1.49 | [1] |
|  | Treatment dosage (mg/kg) | 4.00 | [1, 2] |
| PD | Slope of the effect curve | 4.00 | [1, 2] |

**Table S2.** Pharmacokinetics (PK) and pharmacodynamics (PD) parameter values for drug B that were kept constant throughout the sensitivity analyses.

| **Component** | **Parameter** | **Value** | **Reference** |
| --- | --- | --- | --- |
| **PK** | Absorption rate (per day) | 11.16 | [3] |
|  | Rate at which the drugs move from the central compartment to the peripheral compartment (per day) | 8.46 | [3] |
|  | Rate at which the drugs move from the peripheral compartment to the central compartment (per day) | 3.30 | [3] |
|  | Volume distribution (l/kg) | 173.00 | [3] |
| **PD** | Slope of the effect curve | 6.00 | [2, 3] |

**Figure S10.**
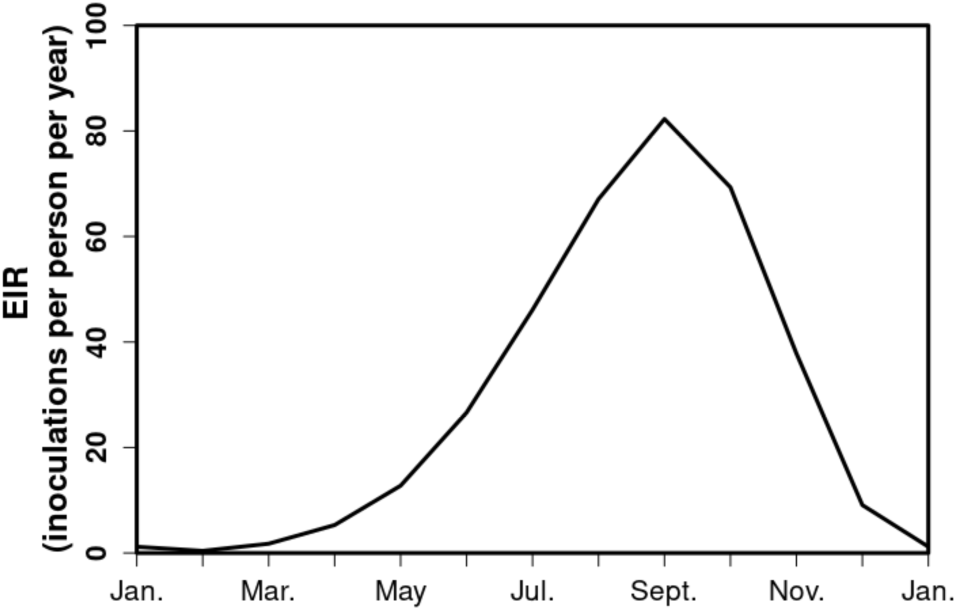
The seasonal transmission of malaria. Example of the EIR (inoculations per person per year) across a year in the seasonal setting of malaria transmission, based on field studies conducted in Tanzania. Here the total EIR is 360 inoculations per person per year.

## 3. Estimation of the selection coefficient

**Figure S11.**
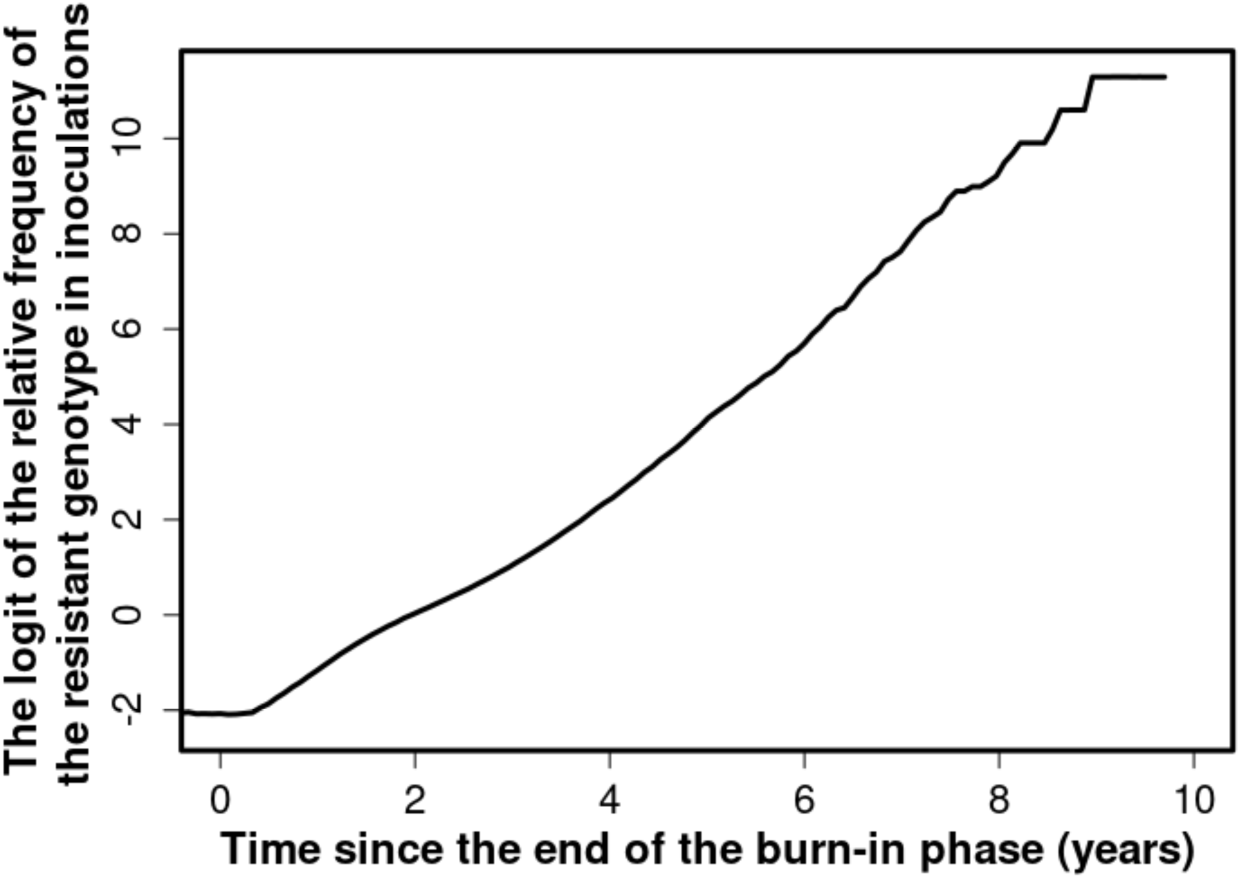
Proof that the selection coefficient is not frequency-*dependent* in OpenMalaria. The figure illustrates the logit of the relative frequency of the resistant genotype over time when the initial relative frequency of infected humans carrying the resistant genotype was 5%. The selection coefficient slope of the logistic regression) was less stable after six years because the percentage of inoculations carrying the sensitive genotype was lower than 0.5%. Thus, the influence of stochastic processes was strong.

**Figure S12.**
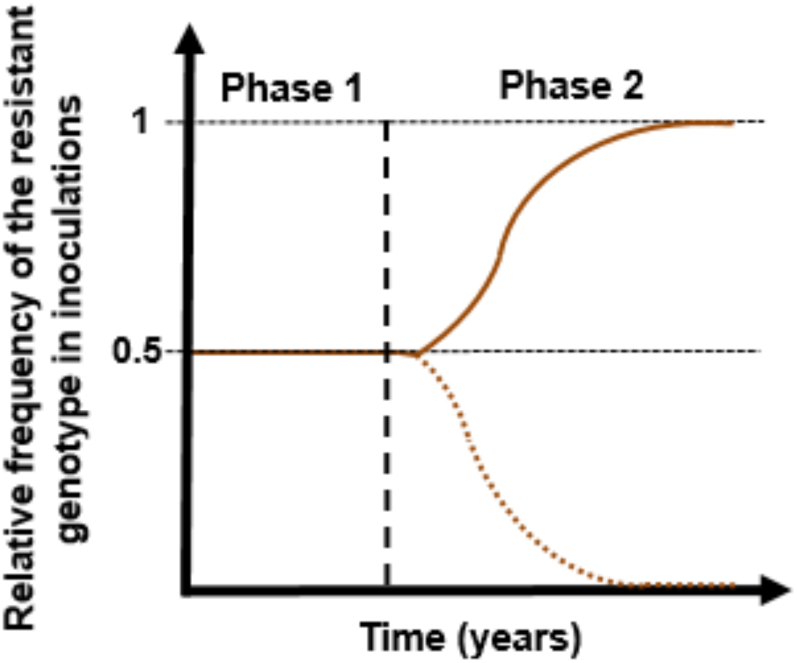
Illustration of typical simulations run in OpenMalaria to estimate the rate of spread of a drug-resistant genotype. The brown line represents the relative frequency of the resistant genotypes in inoculations. The solid line illustrates a simulation in which the resistant genotype spreads in the population (selection coefficient above 0). The dotted line illustrates a simulation in which the resistant genotype did not spread in the population (selection coefficient below 0). Phase 1 represents the burn-in phase. The vertical dotted black line highlights when we introduced the fitness cost and the drug for which the resistant genotype had reduced sensitivity. Phase 2 is the phase during which the rate of spread of the resistant genotype was assessed.

**Figure S13.**
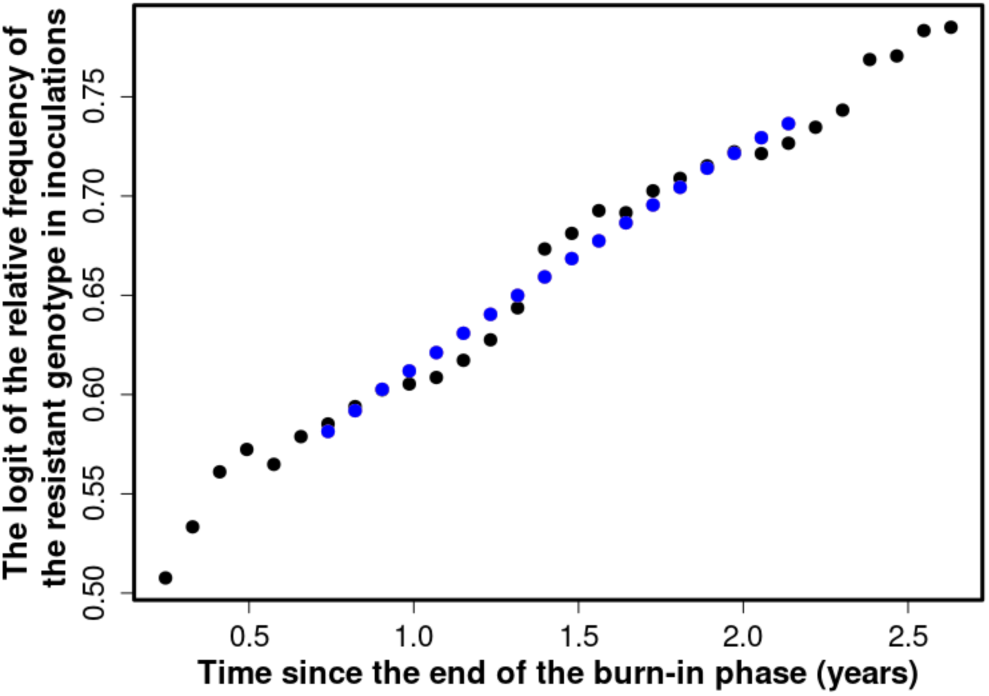
Illustration of the estimation of the selection coefficient in seasonal settings. The black dots represent the logit of the relative frequency of the resistant genotype. The blue dots represent the logit of the moving average of relative frequency of the resistant genotype. The moving average of a measurement at a time t included all the measurements from six months before time t and six months after the time *t*. Using this method, the selection coefficient (slope of the logistic regression) was constant over time.

## 4. Fit of the emulators

**Figure S14.**
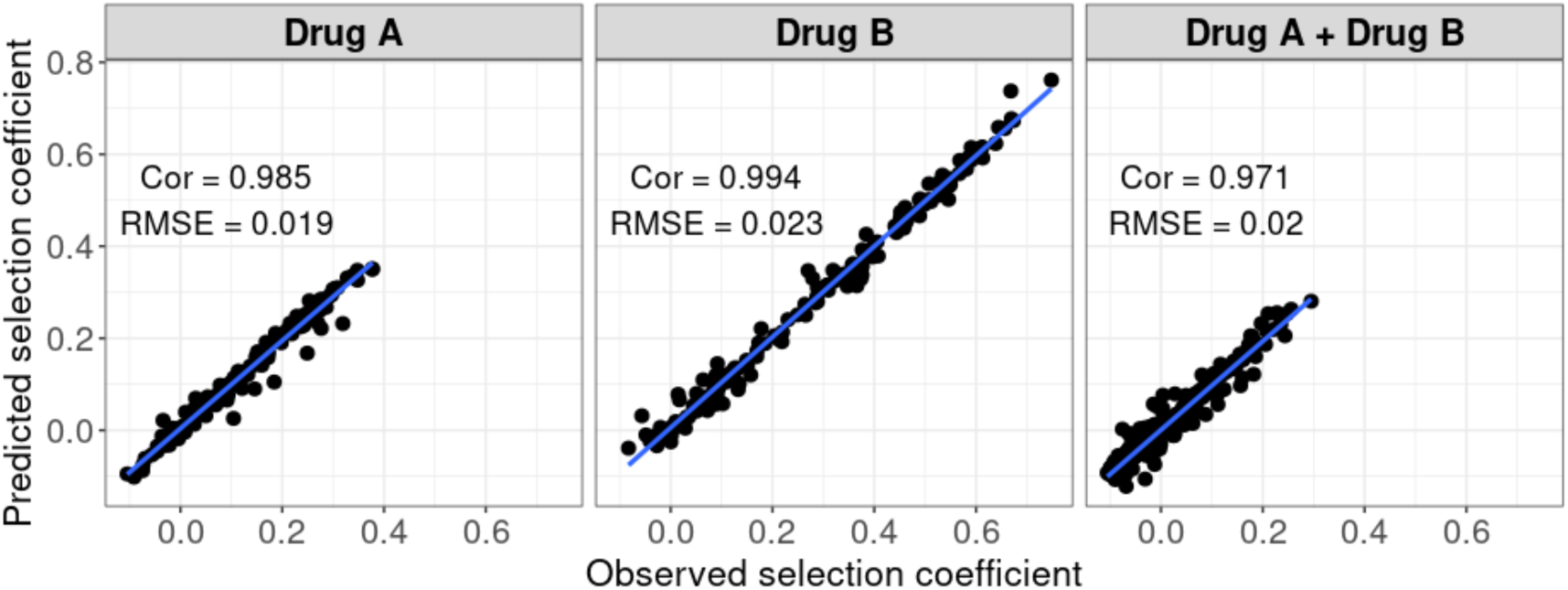
Accuracy of the emulators used for the global sensitivity analyses of each treatment profile. For each treatment profile, the comparison between the selection coefficients of the test dataset estimated using OpenMalaria (i.e., the observed ‘true’ selection coefficient) and the corresponding prediction from the emulator during the final round of adaptive sampling. ‘Cor’ is the Spearman correlation coefficient, ‘RMSE’ is the root means squared error, and the blue lines are the linear regression fits.

**Figure S15.**
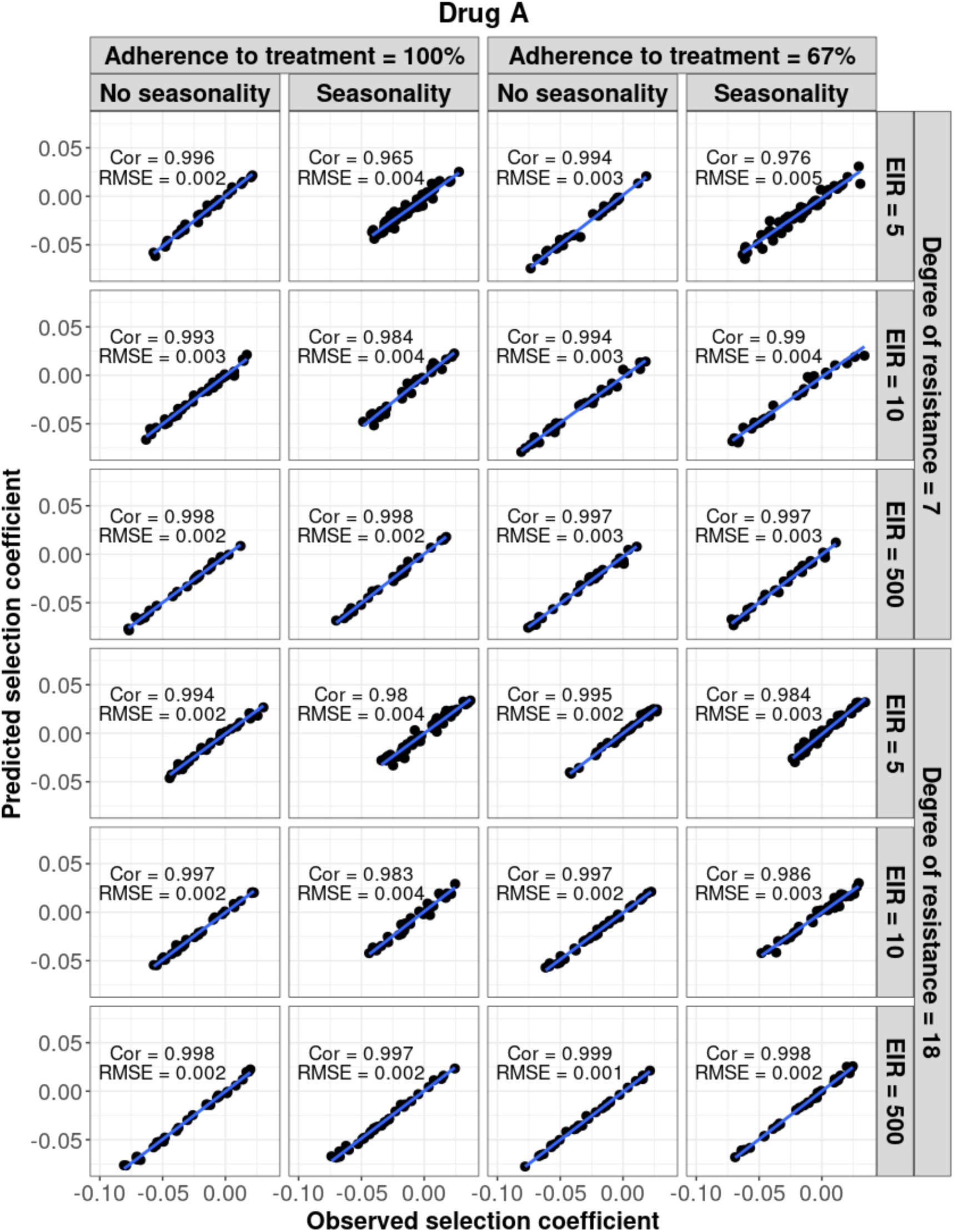
Accuracy of the emulators used for each constrained sensitivity analysis of the spread of a genotype resistant to drug A used in monotherapy in each setting with low access to treatment (10%). The comparison between the selection coefficients for the test dataset between the observed ‘truth’ from OpenMalaria, and the prediction from the emulators during the final round of adaptive sampling. The EIR is in inoculations per person per year (5, 10, and 500). The degree of resistance is the relative decrease in the Emax of the resistant genotype compared with the sensitive one. ‘Cor’ is the Spearman correlation coefficient, ‘RMSE’ is the root means squared error, and the blue lines are the linear regression fits.

**Figure S16.**
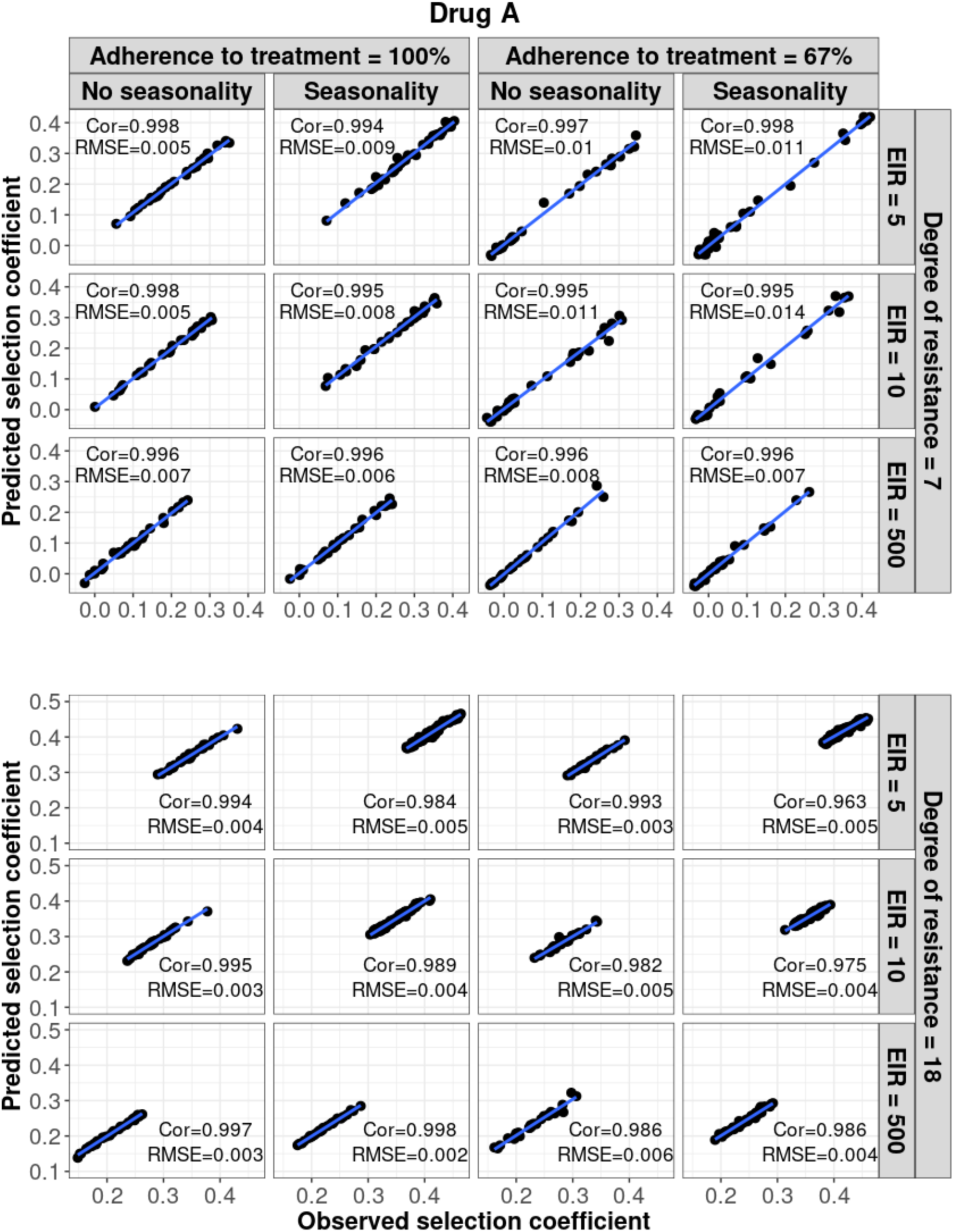
Accuracy of the emulators used for each constrained sensitivity analysis of the spread of a genotype resistant to drug A used in monotherapy in each setting with high access to treatment (80%). The comparison between the selection coefficients for the test dataset between the observed ‘truth’ from OpenMalaria, and the prediction from the emulators during the final round of adaptive sampling. The EIR is in inoculations per person per year (5, 10, or 500). The degree of resistance is the relative decrease in the Emax of the resistant genotype compared with the sensitive one. ‘Cor’ is the Spearman correlation coefficient, ‘RMSE’ is the root means squared error, and the blue lines are the linear regression fits.

**Figure S17.**
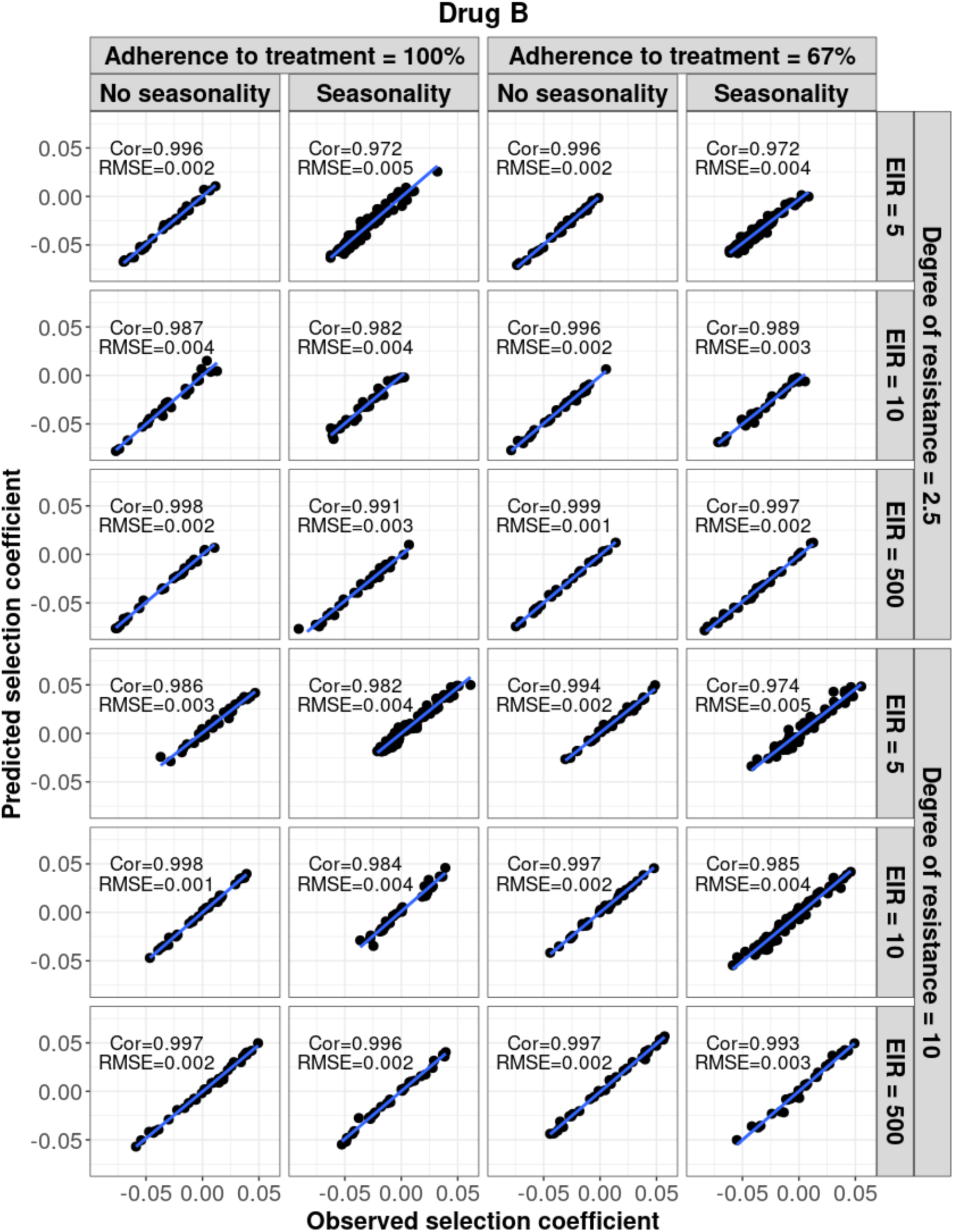
Accuracy of the emulators used for each constrained sensitivity analysis of the spread of a genotype resistant to drug B used in monotherapy in each setting with low access to treatment (10%). The comparison between the selection coefficients for the test dataset between the observed ‘truth’ from OpenMalaria, and the prediction from the emulators during the final round of adaptive sampling. The EIR is in inoculations per person per year (5, 10, or 500). The degree of resistance is the relative increase in the EC50 of the resistant genotype compared with the sensitive one. ‘Cor’ is the Spearman correlation coefficient, ‘RMSE’ is the root means squared error, and the blue lines are the linear regression fits.

**Figure S18.**
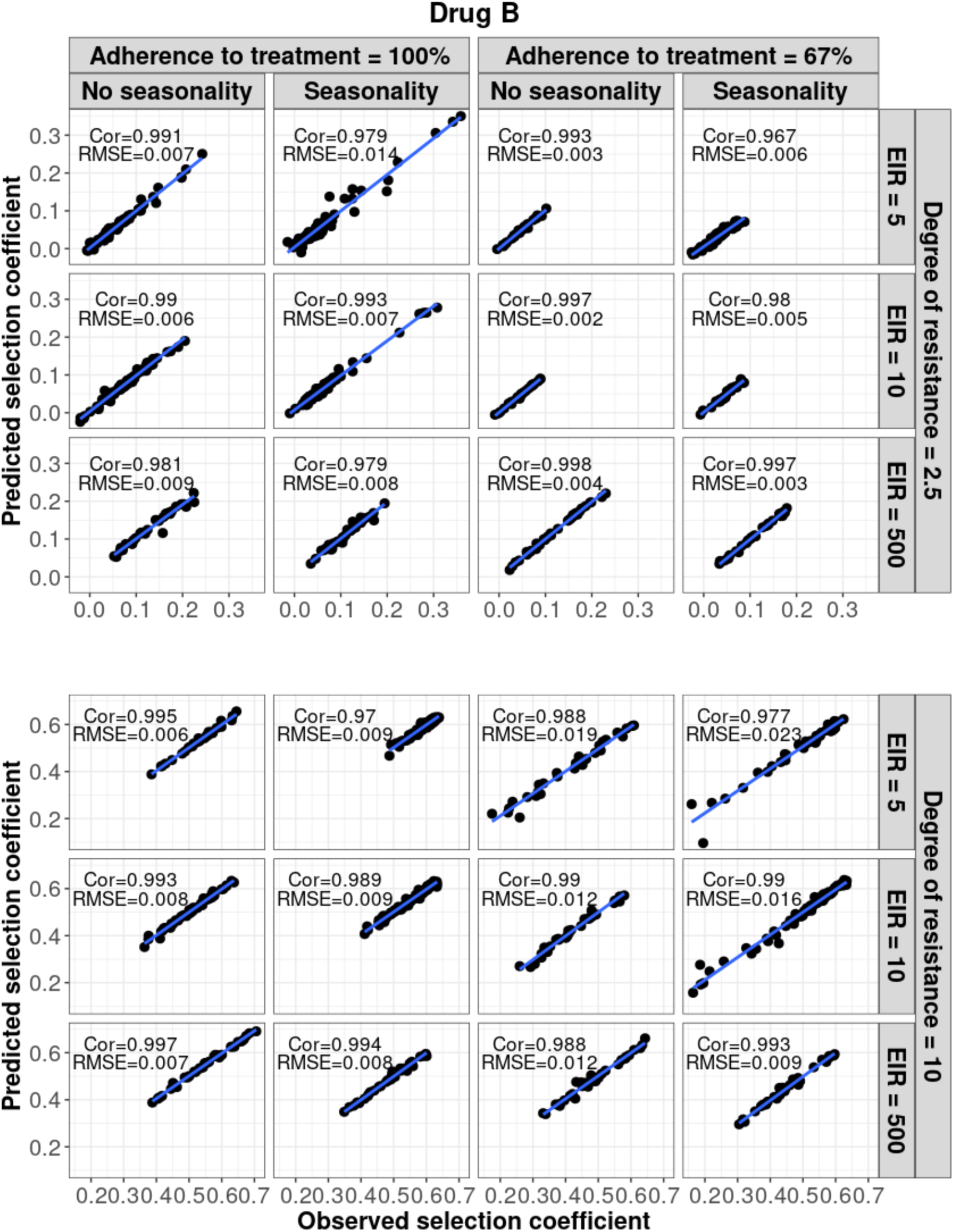
Accuracy of the emulators used for each constrained sensitivity analysis of the spread of a genotype resistant to drug B used in monotherapy in each setting with high access to treatment (80%). The comparison between the selection coefficients for the test dataset between the observed ‘truth’ from OpenMalaria, and the prediction from the emulators during the final round of adaptive sampling. The EIR is in inoculations per person per year (5, 10, or 500). The degree of resistance is the relative increase in the EC50 of the resistant genotype compared with the sensitive one. ‘Cor’ is the Spearman correlation coefficient, ‘RMSE’ is the root means squared error, and the blue lines are the linear regression fits.

**Figure S19.**
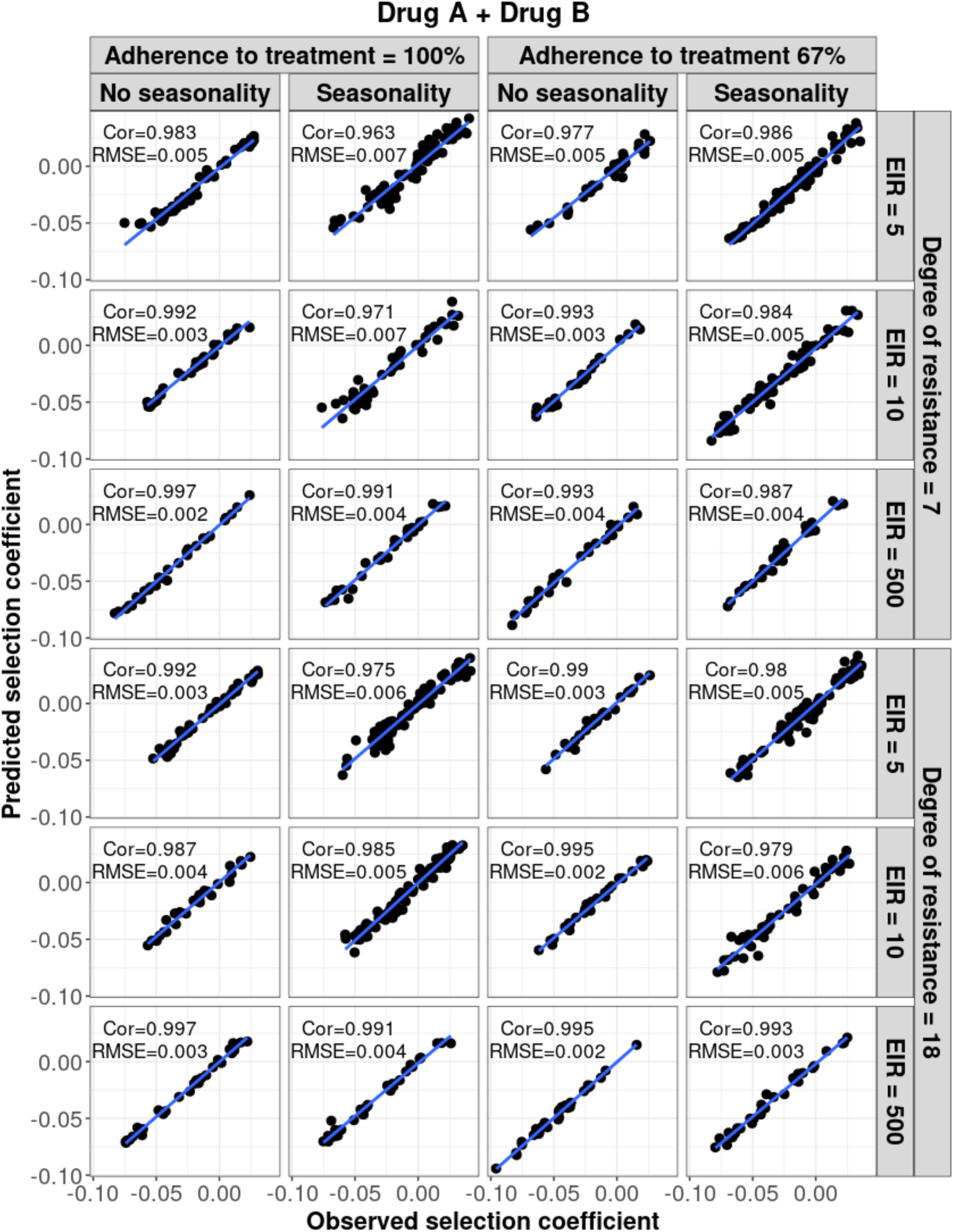
Accuracy of the emulators used for each constrained sensitivity analysis of the spread of a genotype resistant to drug A, when used in combination with drug B, in each setting with low access to treatment (10%). The comparison between the selection coefficients for the test dataset between the observed ‘truth’ from OpenMalaria, and the prediction from the emulators during the final round of adaptive sampling. The EIR is in inoculations per person per year (5, 10, or 500). The degree of resistance to drug A is the relative decrease in the Emax of the resistant genotype compared with the sensitive one. ‘Cor’ is the Spearman correlation coefficient, ‘RMSE’ is the root means squared error, and the blue lines are the linear regression fits.

**Figure S20.**
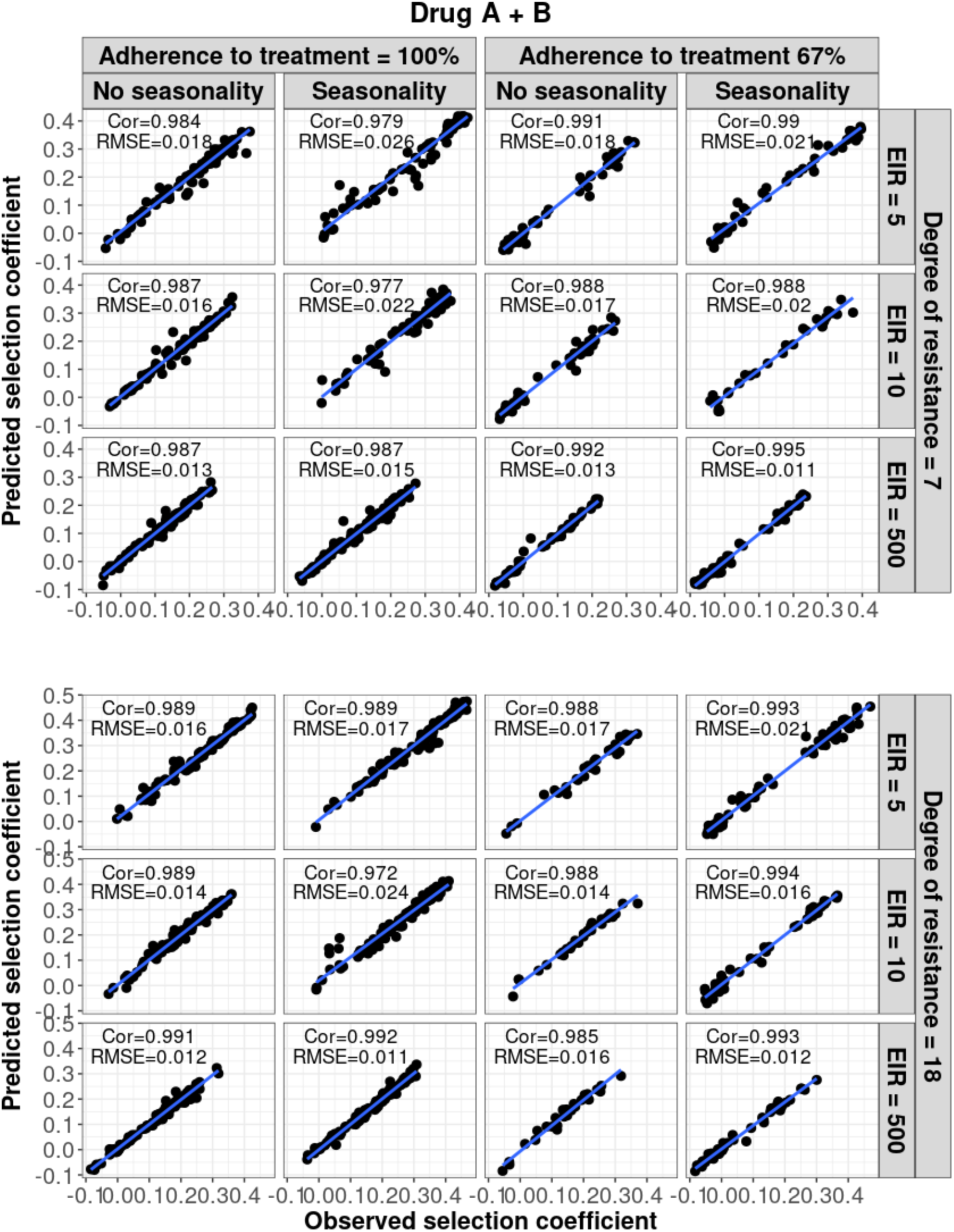
Accuracy of the emulators used for each constrained sensitivity analysis of the spread of a genotype resistant to drug A, when used in combination with drug B, in each setting with high access to treatment (80%). The comparison between the selection coefficients for the test dataset between the observed ‘truth’ from OpenMalaria, and the prediction from the emulators during the final round of adaptive sampling. The EIR is in inoculations per person per year (5, 10, or 500). The degree of resistance to drug A is the relative decrease in the Emax of the resistant genotype compared with the sensitive one. ‘Cor’ is the Spearman correlation coefficient, ‘RMSE’ is the root means squared error, and the blue lines are the linear regression fits.

## 5. Probability of establishment

**Figure S21.**
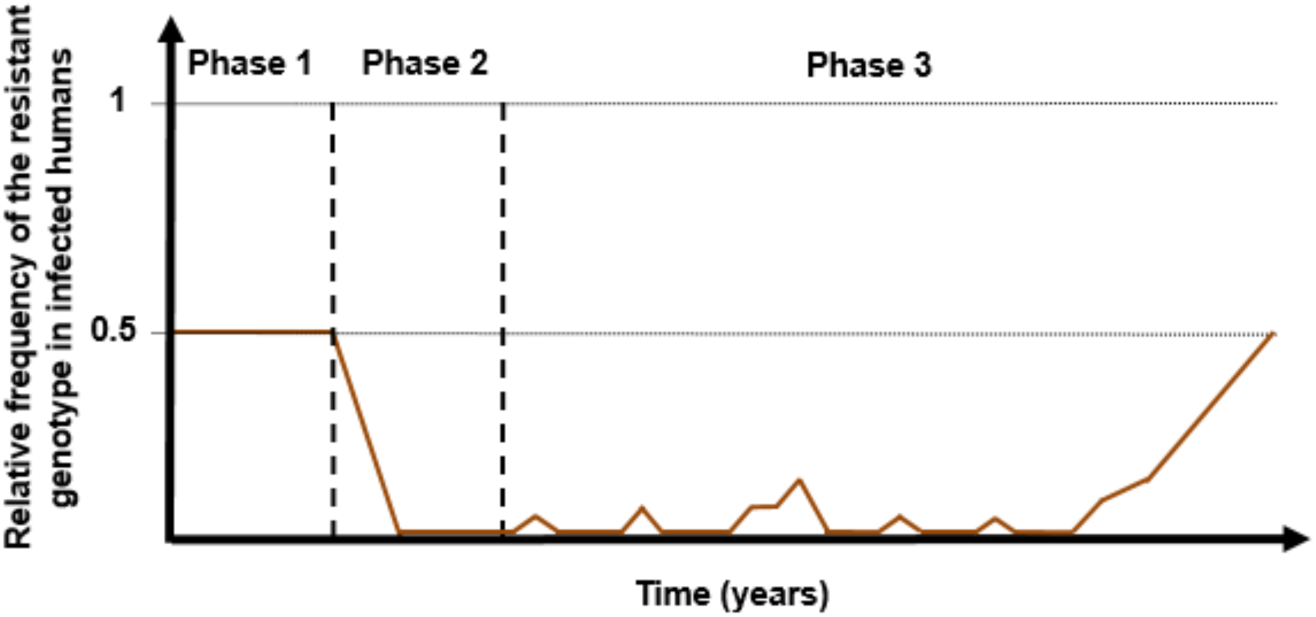
Illustration of typical simulations run in OpenMalaria to estimate the probability of establishment of a drug-resistant genotype. The brown curve represents the relative frequency of the resistant genotypes in inoculations. Phase 1 represents the burn-in phase. In the second phase, we introduced a drug to which resistant parasites were hypersensitive. In the last phase, we imported mutation conferring drug resistance at a low rate until one mutation established.

### 5.1 Calculation of the importation rate for each setting

The importation rate, *I*, (imported infections per 1,000 individuals per year), was calculated to mimic a mutation rate of 5×10^-5^ mutations per infection per year in each setting as in [4]. This low mutation rate ensured that the newly emerged genotype either established or became extinct before a new mutation was imported. The importation rate was calculated as:

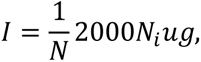

where *N* is the human population size, *N_i_* is the number of infections (i.e., the number of infected people), *u* is the mutation rate per infection (i.e., per transmission), and *g* is the number of malaria generations per year. Thus, *N_i_ug* represents the number of *de novo* resistant mutations transmitted per year. This number was divided by the human population size and multiplied by 1,000 to obtain the number of imported infections per 1,000 persons per year. This is multiplied by two, as half of the imported infections were sensitive.

